# Effectiveness of an Inactivated Covid-19 Vaccine with Homologous and Heterologous Boosters against Omicron in Brazil

**DOI:** 10.1101/2022.03.30.22273193

**Authors:** Otavio T. Ranzani, Matt D.T. Hitchings, Rosana Leite de Melo, Giovanny V. A. de França, Cássia de Fátima R. Fernandes, Margaret L Lind, Mario Sergio Scaramuzzini Torres, Daniel Henrique Tsuha, Leticia C. S. David, Rodrigo F. C. Said, Maria Almiron, Roberto D. de Oliveira, Derek A.T. Cummings, Natalie E Dean, Jason R. Andrews, Albert I. Ko, Julio Croda

**Author notes:** Correspondence to: Prof Julio Croda, Universidade Federal de Mato Grosso do Sul and Fundação Oswaldo Cruz. Authors contributed equally (Derek A.T. Cummings, Natalie E Dean, Jason R. Andrews, Albert I. Ko, Julio Croda).

## Abstract

The effectiveness of inactivated vaccines (VE) against symptomatic and severe COVID-19 caused by omicron is unknown. We conducted a nationwide, test-negative, case-control study to estimate VE for homologous and heterologous (BNT162b2) booster doses in adults who received two doses of CoronaVac in Brazil in the Omicron context. Analyzing 1,386,544 matched-pairs, VE against symptomatic disease was 8.6% (95% CI, 5.6-11.5) and 56.8% (95% CI, 56.3-57.3) in the period 8-59 days after receiving a homologous and heterologous booster, respectively. During the same interval, VE against severe Covid-19 was 73.6% (95% CI, 63.9-80.7) and 86.0% (95% CI, 84.5-87.4) after receiving a homologous and heterologous booster, respectively. Waning against severe Covid-19 after 120 days was only observed after a homologous booster. Heterologous booster might be preferable to individuals with completed primary series inactivated vaccine.

## Introduction

The substantial initial protection of primary series Covid-19 vaccines against moderate and severe Covid-19 has been demonstrated through randomized clinical trials and observational studies.^1–3^ Since then, accumulating evidence has demonstrated the importance of waning protection following primary series completion,^4–6^ and decreased effectiveness of current vaccines to variants of concern (VoC), in particular the Omicron (B.1.1.529) variant.^4,7^ Delineating the effectiveness of the range of booster vaccination strategies is therefore critical for guiding national and global policy.^8^

The majority of the existing vaccine effectiveness evidence is for mRNA vaccines and adenoviral vectored vaccines, both as the primary series and as booster doses,^7,9,10^ leaving significant evidence gaps regarding inactivated vaccine products. Inactivated vaccines are widely used, particularly in low- and middle-income countries, and represent half of the administered doses of Covid-19 vaccines worldwide as of Jan 2022.^11^ Large Omicron epidemics associated with severe cases and deaths have occurred in regions, most recently Eastern Asia, where inactivated vaccines have been extensively administered.^12^ Brazil initiated booster vaccination in September 2021, after Delta VoC began to dominate in the country and three months before Omicron dominance.^5^ Evidence concerning the effectiveness of inactivated vaccines with homologous or heterologous boosters is critically needed to inform vaccine policies in countries that used these vaccines in their initial rollout.

We evaluated the vaccine effectiveness of CoronaVac and BNT162b2 booster doses among Brazilian adults who completed the primary series of the CoronaVac vaccine in a nationwide test-negative case-control study. Our primary analysis focused on the period from December 25, 2021 to April 22, 2022, when circulation of the Omicron variant was predominant, and compared these findings with those from the prior period, from September 6, 2021 to December 14, 2021 when the Delta variant was predominant in the country.

## Methods

### Study setting and design

We conducted a matched test-negative case-control study between September 6, 2021, and April 22, 2022, in Brazil. The national Covid-19 vaccination campaign started on January 17, 2021, and administration of booster doses began for the general population on September 6, 2021. The primary series used in Brazil were homologous schemes of Sinovac CoronaVac (two doses), Oxford-AstraZeneca ChAdOx1 nCoV-19 (two doses), Pfizer BNT162b2 (two doses), Janssen Ad26.COV2.S (single dose), and heterologous combinations of the above products in periods of vaccine shortage. All four vaccine products were administered as a homologous or heterologous booster dose. There was no differential recommendation for which vaccine to be administered, except a suggestion for BNT162b2 if available. The booster vaccination followed an age-prioritization scheme. The interval between second and booster doses was initially six months and was subsequently shortened to four months during November 2021 in some states and nationally on December 20, 2021. The proportion of individuals with a primary series of CoronaVac who received a booster dose of Ad26.COV2.S or ChAdOx1 nCoV-19 was small; therefore we limited our analysis to booster doses of CoronaVac and BNT162b2.

### Data sources

We obtained individual-level information on Covid-19 outcomes from two national surveillance databases in Brazil: e-SUS and SIVEP-Gripe. e-SUS collects information of any individual suspected to have mild Covid-19 syndromic illnesses, including those who were not tested, tested negative and tested positive. SIVEP-Gripe collects information on any severe acute respiratory infection, including all Covid-19 hospitalizations and deaths.^3,5,13^ We obtained individual-level vaccination status from the national vaccination database (SI-PNI). Notification to these three systems is compulsory in Brazil. The three databases have a unique identifier after pseudo-anonymization by the Ministry of Health. Additional information is available on **eTable 1**. We extracted eSUS, SIVEP-Gripe and SI-PNI on 29/04/2022 and used data until 22/04/2022, allowing for a one-week potential delay. This study was approved by the ethical committee for research of Federal University of Mato Grosso do Sul (CAAE: 43289221.5.0000.0021)

The study population was adults (aged ≥18 years) residing in Brazil, and who underwent SARS-CoV-2 RT-PCR or rapid antigen testing associated with symptomatic illness^14^ during the study period. We excluded individuals with missing or inconsistent information on age, sex, municipality of residence, and on vaccination and testing status and dates. We excluded RT-PCR/antigen tests that were not collected within 10 days of symptom onset to avoid potentially misclassification, positive or negative RT-PCR/antigen tests with a positive RT-PCR/antigen test in the previous 90 days to capture only incident infections and avoid a second positive test because of prolonged viral shedding, and negative RT-PCR/antigen tests with a positive RT-PCR/antigen test occurring in the following 14 days because of likely false-negative test in the first negative test. For individuals who received multiple RT-PCR or antigen tests during the study period, we included all eligible tests up to and including the first positive test (ie, the first positive test in the study period and at least 90 days prior to another positive). The number of RT-PCR/antigen tests performed during the study period in Brazil is shown in **eFigure 1**.

**Figure 1.**
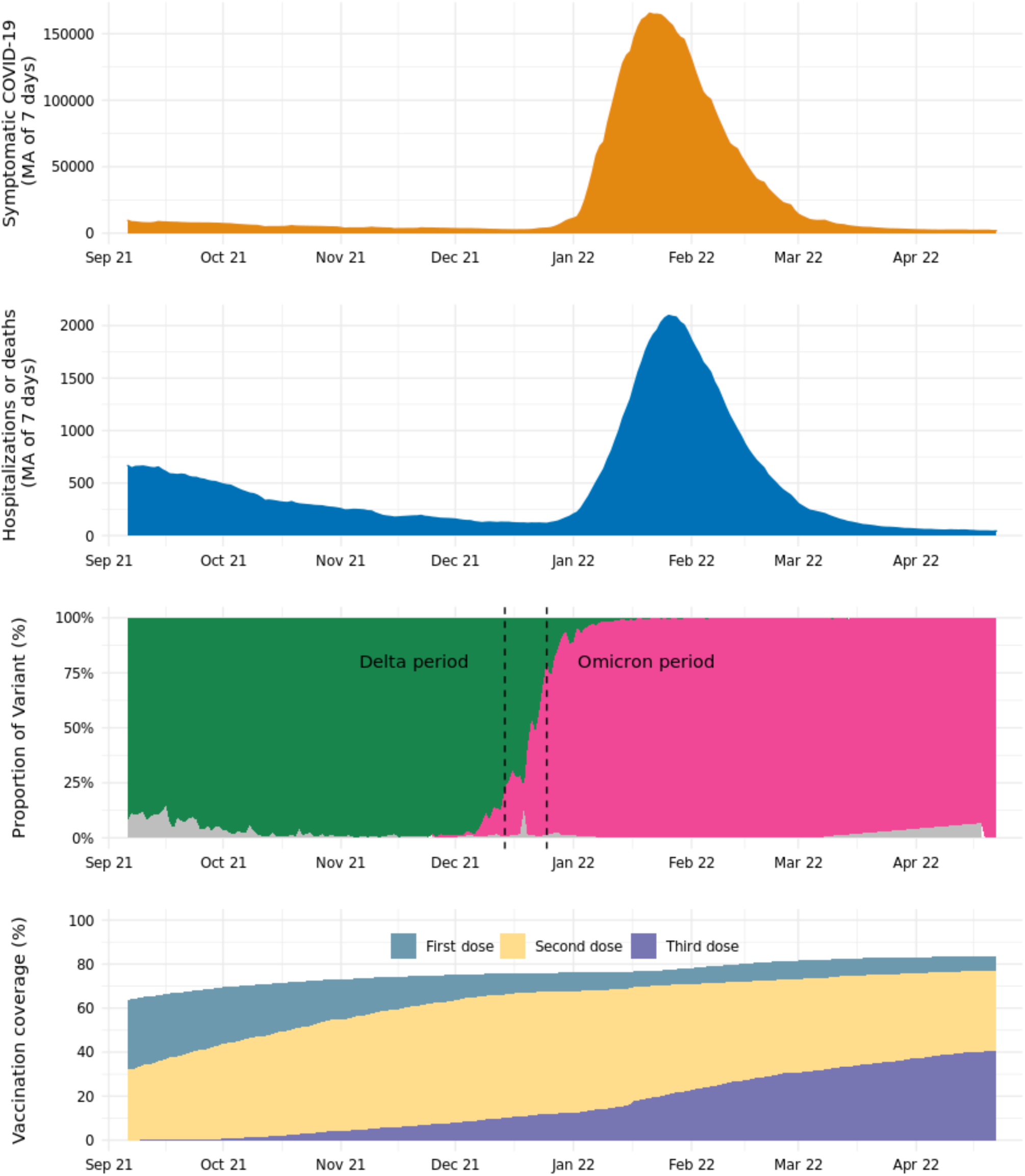
Times series of Covid-19 cases and Covid-19 hospital admissions or deaths, variants of concern prevalence and vaccination coverage in Brazil from Sep 2021 to Apr 2022. Daily prevalence of SARS-CoV-2 variants among genotyped isolates were obtained from the GISAID (global initiative on sharing avian influenza data) database (extraction on 09 May 2022), selecting samples from Brazil. Green represents Delta prevalence, pink area represents Omicron prevalence and gray area represents others. Second dose coverage includes a single dose of Ad26.COV2.S. MA - moving averages.

To assess waning of the booster doses over time since administration, we performed a separate, secondary, case-control analysis on the same study population, restricting to cases and controls who received a primary series of CoronaVac and received an RT-PCR/antigen test at least six months after their second dose, i.e. when eligible for a booster dose. The study design and matching procedure was otherwise the same.

### Selection of cases and matched controls

Cases were defined as those from the study population who had Covid-19 symptoms, defined by the presence of at least one symptom: fever, sore throat, headache, cough, chills, runny nose, dyspnea, anosmia, and ageusia, and a positive SARS-CoV-2 RT-PCR/antigen test result. Eligible controls were defined as those from the study population who had Covid-19 symptoms as defined by cases, and a negative SARS-CoV-2 RT-PCR/antigen test result. Finally, we excluded all RT-PCR/antigen tests that were obtained after receipt of a primary series of ChAdOx1 nCoV-19, BNT162b2 or Ad26.COV2.S vaccines.

We matched each case with one control according to the age (± 10 years), sex, municipality of residence, variant period, and RT-PCR/antigen test sample collection date (± 10 days). The algorithm used for the continuous variables (age and test sample collection date) was nearest neighbor matching. After identification of each case, we randomly chose one control from the set of all eligible matching controls, allowing for replacement of controls. We performed a sensitivity analysis on the matching approach by creating strata of unique combinations of the matching factors (age category in 10 years band, sex, municipality of residence, variant period and week of testing). The numbers of cases and controls per stratum are not pre-specified, and strata with no cases, or with no controls, were excluded. This leads to varying ratios of cases to controls. This was done to use all available information, reducing how often unmatched cases or controls needed to be discarded and no case or control appears in more than one stratum, thus dealing with the potential issue related to replacement. To improve computational performance of the models while retaining the majority of cases, large strata were reduced in size by dividing into smaller strata. In strata with more controls than cases, each stratum allowed a case to be matched to up to ten controls, and vice versa. For strata with at least ten times as many controls as cases, excess controls were discarded, and vice versa. In this way, strata sizes varied from two to eleven.^15^

### Statistical analysis

We estimated the vaccine effectiveness of booster doses of CoronaVac and BNT162b2 against symptomatic Covid-19 in the 0-7 days, 8-59 days and ≥60 days after the booster dose. We also estimated the vaccine effectiveness of a booster dose against Covid-19 hospitalization and/or death by restricting the analysis population to case-control pairs in which the case was hospitalized or died.^3,5,16,17^ Symptomatic Covid-19 includes mild and severe cases. Severe Covid-19 was defined as hospital admission and death with severe acute respiratory infection due to SARS-CoV-2 (positive RT-PCR/Antigen test). For the analyses of symptomatic and severe Covid-19, we considered the date of respiratory sample collection as the date of the event. There are several choices of controls for severe outcomes, including community non-syndromic controls, community syndromic controls, and hospitalized test-negative controls with or without symptoms.^16,18,19^ Each has their advantages and disadvantages in how well they represent the source population in their uptake of Covid-19 vaccination. We choose a priori to use community and hospitalized syndromic controls as we agreed these are the controls that better represent the vaccination status in the Brazilian setting. Additionally, we chose syndromic controls to reduce the bias in testing behaviour,^20^ as those tested in the absence of symptoms are more likely to be part of special groups of individuals (e.g., healthcare workers). The reference group was unvaccinated individuals. For the secondary analysis assessing waning effectiveness, we estimated the relative vaccine effectiveness (rVE),^7,21^ using booster eligible (≥180 days after the second dose) CoronaVac recipients as the reference group, and stratified the time since booster administration by 8-59 days, 60-89 days, 90-119 days and ≥120 days. We used RT-PCR/Antigen test respiratory samples to define cases and controls in any effectiveness analyses. Cases and controls that were not linked to the vaccination database were ascertained as unvaccinated.

We used conditional logistic regression to estimate the adjusted odds ratio (aOR) of vaccination comparing cases and controls, and (1−aOR)*100 provided an estimate of vaccine effectiveness under the assumptions of a test negative design.^16^ Because age is a strong determinant of Covid-19 outcomes, we adjusted for age (as a continuous variable, modeled with a restricted cubic spline) after matching to control for potential residual confounding within age bands. We also adjusted for chronic comorbidities (including cardiovascular, renal, diabetes, chronic respiratory disorder, obesity, or immunosuppression, categorized as 0, 1, and ≥2 comorbidities), self-reported race, and any previous symptomatic event that were reported to the surveillance systems (categorized as 0, and ≥1). Prior SARS-CoV-2 exposure is defined as notified acute respiratory infection or positive SARS-CoV-2 test result prior to the sampled RT-PCR/Antigen test. This variable is our best surrogate of previously confirmed or suspected infection of SARS-CoV-2. We considered the vaccine effectiveness estimates for the 0-13 days after the first dose as a “bias indicator”, because it is expected that vaccines have no or limited effectiveness for this period.^22^

We conducted an analysis of vaccine effectiveness within age subgroups (<60, 60-74 and vs ≥75 years old) by adding an interaction term with the vaccination category. Because the analysis period incorporated a Delta (B.1.617.2) (September 6, 2021 to December 14, 2021) and Omicron (December 25, 2021 to April 22, 2022) period, we conducted separate analyses in each time period. We defined the end of the Delta period as when national Omicron VoC prevalence amongst sequenced genomes reached 25% and the beginning of the Omicron period as when the prevalence reached 75% in the GISAID database.^23^ We conducted the same analyses using only RT-PCR tests as a sensitivity analysis, to address potential misclassification. Finally, in a post-hoc sensitivity analysis, we evaluated vaccine effectiveness in the main analysis population further adjusting by month of second dose as a factor in the model.

All analyses were done in R (v.4.1.2).^24^

## Results

### Descriptive Characteristics

During the study period, there was a low incidence of Covid-19 cases and hospital admissions or deaths during the Delta wave compared to earlier periods in Brazil, until the end of December 2021, which corresponded to the introduction and spread of the Omicron variant (**Figure 1**). During Omicron period, 97.3% of samples were BA.1 (97.3%) followed by BA.2 (2.7%) according to the GISAID data. Across all age groups in Brazil, on April 22, 2022, coverage was 83.7% for the first vaccine dose, 77.1% for second doses, and 40.7% for boosters (**Figure 1**).

After applying the inclusion and exclusion criteria, there were 3,548,209 RT-PCR/antigen tests from 3,320,429 individuals eligible for matching for the primary analysis. After matching one control per case, with replacement, the analysis population was 3,094,478 RT-PCR/antigen tests from 2,107,696 individuals for the primary analysis (**eFigure 2**). Controls were matched to multiple cases a mean 2.7 ± 4, median of 2 (IQR: 1-3) times. The characteristics for the selected case-control sets for CoronaVac during the Delta and Omicron periods is shown in **Table 1**. Among severe Covid-19, 88% (54,307/61,647) of patients had at least one sign/symptom of respiratory distress (dyspnoea, hypoxaemia, respiratory discomfort); 77% (46,739/60,919) of patients received either non-invasive or invasive mechanical ventilation. During the omicron period, these numbers were 87% (36,702/42,195) and 75% (31,151/41,714) respectively. The characteristics of those who received an homologous or heterologous booster are shown on **eTable 2**.

**Figure 2.**
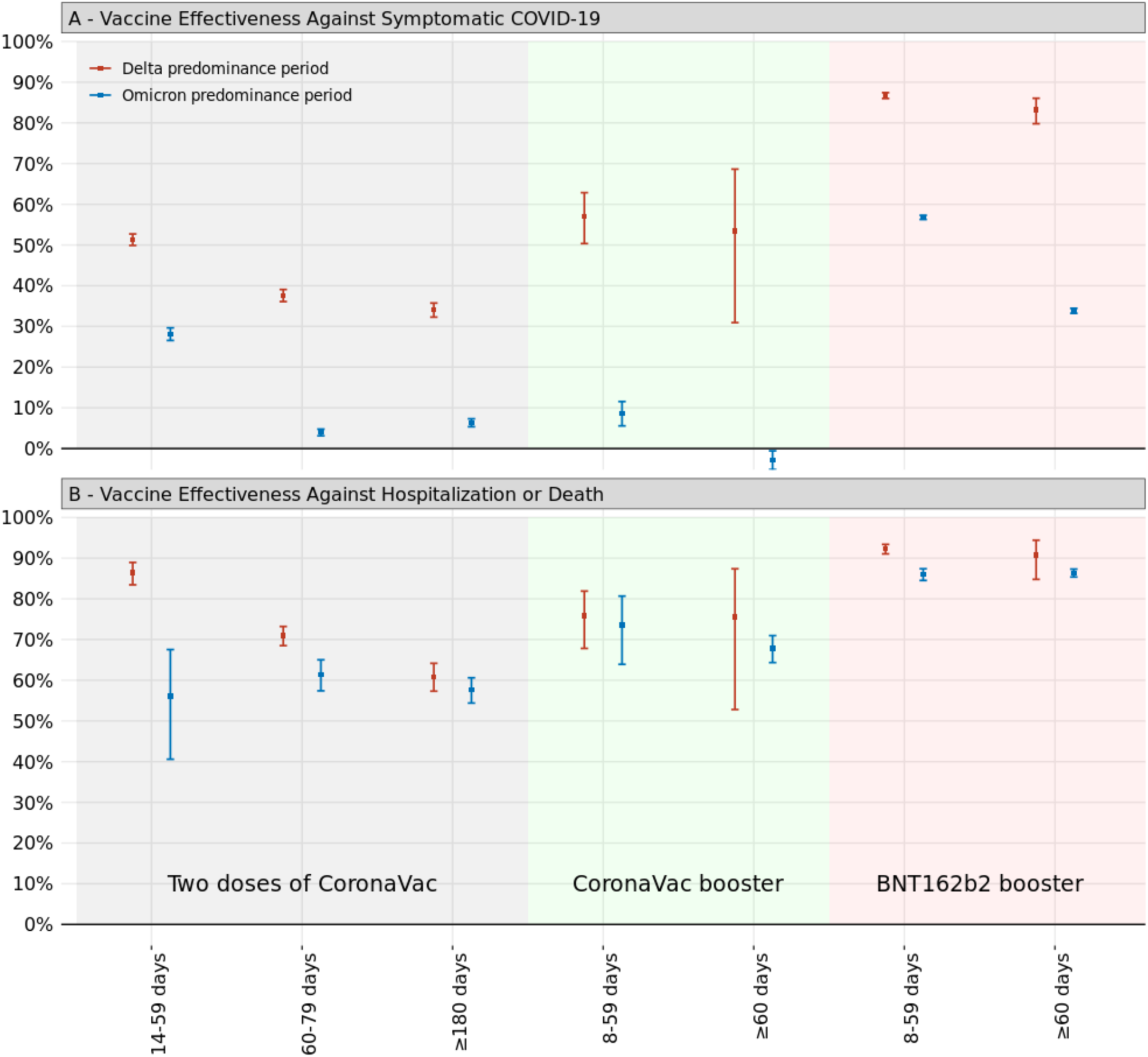
Adjusted vaccine effectiveness against symptomatic Covid-19 (A) and Covid-19 hospitalization or death (B) of primary two-dose vaccination with CoronaVac and subsequent booster vaccination with CoronaVac and BNT162b2 compared to unvaccinated individuals, according to days since receiving the last vaccine dose.

**Table 1.** Characteristics of adults in Brazil, who were selected into case test negative pairs for the analysis of vaccine effectiveness during the Delta period (September 6, 2021 to December 14, 2021) and the Omicron period (December 25, 2021 to Apr 22, 2022)

|  | Matched pairs for Delta period |  |  | Matched pairs for Omicron period |  |  |
| --- | --- | --- | --- | --- | --- | --- |
|  | Controls<br>(n=160,695) | Cases<br>(n=160,695) | SMD | Controls<br>(n=1,386,544) | Cases<br>(n=1,386,544) | SMD |
| **Demographics** |  |  |  |  |  |  |
| **Age, mean (SD), years** | 45.3 (19.7) | 45.5 (19.8) | 0.011 | 42.6 (18.7) | 42.9 (18.9) | 0.019 |
| **Age categories, n (%)** |  |  | 0.039 |  |  | 0.052 |
| 18-39 years | 80624 (50.2) | 78822 (49.1) |  | 788973 (56.9) | 770276 (55.6) |  |
| 40-59 years | 33251 (20.7) | 35150 (21.9) |  | 279260 (20.1) | 297263 (21.4) |  |
| 60-79 years | 40746 (25.4) | 39948 (24.9) |  | 276213 (19.9) | 267538 (19.3) |  |
| ≥80 years | 6074 ( 3.8) | 6775 ( 4.2) |  | 42098 ( 3.0) | 51467 ( 3.7) |  |
| **Male sex, n (%)** | 74312 (46.2) | 74312 (46.2) | <0.001 | 576604 (41.6) | 576604 (41.6) | <0.001 |
| **Self-reported race<sup>†</sup>, n (%)** |  |  | 0.029 |  |  | 0.052 |
| White/Branca | 68661 (42.7) | 69853 (43.5) |  | 604233 (43.6) | 620750 (44.8) |  |
| Brown/Pardo | 50272 (31.3) | 48671 (30.3) |  | 423463 (30.5) | 405572 (29.3) |  |
| Black/Preta | 6304 ( 3.9) | 6062 ( 3.8) |  | 57866 ( 4.2) | 51664 ( 3.7) |  |
| Yellow/ Amarela | 1969 ( 1.2) | 2203 ( 1.4) |  | 25547 ( 1.8) | 23496 ( 1.7) |  |
| Indigenous/Indigena | 1171 ( 0.7) | 1358 ( 0.8) |  | 5189 ( 0.4) | 2983 ( 0.2) |  |
| Missing | 32318 (20.1) | 32548 (20.3) |  | 270246 (19.5) | 282079 (20.3) |  |
| **Region of residence** |  |  | <0.001 |  |  | <0.001 |
| North | 11901 ( 7.4) | 11901 ( 7.4) |  | 67977 ( 4.9) | 67977 ( 4.9) |  |
| Northeast | 21128 (13.1) | 21128 (13.1) |  | 200629 (14.5) | 200629 (14.5) |  |
| Central-West | 14865 ( 9.3) | 14865 ( 9.3) |  | 108722 ( 7.8) | 108722 ( 7.8) |  |
| Southeast | 73931 (46.0) | 73931 (46.0) |  | 743956 (53.7) | 743956 (53.7) |  |
| South | 38870 (24.2) | 38870 (24.2) |  | 265260 (19.1) | 265260 (19.1) |  |
| **Reported number of chronic comorbidities<sup>‡</sup>, n (%)** |  |  | 0.058 |  |  | 0.012 |
| None | 141535 (88.1) | 138508 (86.2) |  | 1281876 (92.5) | 1285339 (92.7) |  |
| One or two | 18254 (11.4) | 20972 (13.1) |  | 101645 ( 7.3) | 97797 ( 7.1) |  |
| Three or more | 906 ( 0.6) | 1215 ( 0.8) |  | 3023 ( 0.2) | 3408 ( 0.2) |  |
| **Prior SARS-CoV-2 exposure** |  |  |  |  |  |  |
| Previous symptomatic events notified to the surveillance system<sup>¶</sup>, n (%) | 41632 (25.9) | 28920 (18.0) | 0.192 | 336835 (24.3) | 345715 (24.9) | 0.015 |
| Positive SARS-CoV-2 test result<sup>††</sup>, n (%) | 9159 ( 5.7) | 2594 ( 1.6) | 0.219 | 92402 ( 6.7) | 66049 ( 4.8) | 0.082 |
| **Interval between symptoms onset and RT-PCR/Antigen testing, median (p25-p75), days** | 3 [2, 5] | 3 [2,5] | 0.064 | 3 [2,4] | 3 [2,4] | 0.140 |
| **Hospitalization or death** | 7910 ( 4.9) | 18922 (11.8) | 0.250 | 22307 ( 1.6) | 35807 ( 2.6) | 0.252 |
| **Vaccination status** |  |  | 0.313 |  |  | 0.206 |
| Not vaccinated, n (%) | 36058 (22.4) | 50002 (31.1) |  | 165850 (12.0) | 197214 (14.2) |  |
| *Primary vaccination with CoronaVac* |  |  |  |  |  |  |
| Single dose, 0-13 days, n (%) | 2104 ( 1.3) | 2606 ( 1.6) |  | 1705 ( 0.1) | 1472 ( 0.1) |  |
| Single dose, ≥14 days, n (%) | 13330 ( 8.3) | 13497 ( 8.4) |  | 74669 ( 5.4) | 71579 ( 5.2) |  |
| Two doses, 0-13 days, n (%) | 4291 ( 2.7) | 3678 ( 2.3) |  | 3306 ( 0.2) | 2991 ( 0.2) |  |
| Two doses, 14-89 days, n (%) | 30019 (18.7) | 24131 (15.0) |  | 57216 ( 4.1) | 50099 ( 3.6) |  |
| Two doses, 90-179 days, n (%) | 37211 (23.2) | 35791 (22.3) |  | 385196 (27.8) | 431676 (31.1) |  |
| Two doses, ≥180 days, n (%) | 24162 (15.0) | 25515 (15.9) |  | 134379 ( 9.7) | 161176 (11.6) |  |
| *Booster vaccination* |  |  |  |  |  |  |
| Third dose of CoronaVac, 0-7 days, n (%) | 104 ( 0.1) | 103 ( 0.1) |  | 1111 ( 0.1) | 1170 ( 0.1) |  |
| Third dose of CoronaVac, 8-59 days, n (%) | 539 ( 0.3) | 466 ( 0.3) |  | 7666 ( 0.6) | 8589 ( 0.6) |  |
| Third dose of CoronaVac, ≥60 days, n (%) | 71 ( 0.0) | 81 ( 0.1) |  | 21364 ( 1.5) | 27847 ( 2.0) |  |
| Third dose of BNT162b2, 0-7 days, n (%) | 1977 ( 1.2) | 1945 ( 1.2) |  | 18549 ( 1.3) | 18486 ( 1.3) |  |
| Third dose of BNT162b2, 8-59 days, n (%) | 10259 ( 6.4) | 2654 ( 1.7) |  | 137230 ( 9.9) | 75833 ( 5.5) |  |
| Third dose of BNT162b2, ≥60 days, n (%) | 570 ( 0.4) | 226 ( 0.1) |  | 378303 (27.3) | 338412 (24.4) |  |
| **Interval between first dose and testing, median (p25-p75), days** | 33 [20, 63] | 33 [19, 63] | 0.004 | 137 [93, 166] | 141 [99, 171] | 0.068 |
| **Interval between second dose and testing, median (p25-p75), days** | 140 [58, 180] | 147 [70, 186] | 0.109 | 139 [117, 174] | 142 [119, 179] | 0.061 |
| **Interval between third dose and testing, median (p25-p75), days** | 22 [11, 38] | 10 [6, 30] | 0.325 | 86 [51, 105] | 91 [64, 109] | 0.177 |

RT-PCR=reverse transcription polymerase chain reaction; SMD=standardized mean difference; SD=standard deviation; † Race/skin colour as defined by the Brazilian national census bureau (Instituto Nacional de Geografia e Estatísticas). ‡ Comorbidities included cardiovascular, or renal conditions, diabetes, chronic respiratory disorder, obesity, or immunosuppression. ¶ Reported illness with covid-19 associated symptoms in eSUS and SIVEP-Gripe databases before the start of study on 06 September 2021.. \*\* Defined as a positive SARS-CoV-2 RT-PCR or antigen detection test result before the start of study on 06 September 2021.

### Vaccine effectiveness estimates

Vaccine effectiveness estimates for two doses of CoronaVac, and for a booster dose of CoronaVac and BNT162b2, are displayed in **Figure 2** and in **eTable 3**. Relative to the Delta period, the Omicron period was associated with a substantial decrease in vaccine effectiveness against symptomatic disease for the primary series of CoronaVac (VE ≥180 days after second dose 34.0%, 95% CI 32.3 to 35.7; in the Delta period; compared to 6.3%, 95% CI 5.3 to 7.3, during the Omicron period). During the Omicron period, vaccine effectiveness 8-59 days after a homologous booster was 8.6% (95% CI, 5.6 to 11.5) against symptomatic Covid-19 and 73.6% (95% CI, 63.9 to 80.7) against severe Covid-19 and for a BNT162b2 booster, vaccine effectiveness was 56.8% (95% CI, 56.3 to 57.3) against symptomatic and 86.0% (95% CI, 84.5 to 87.4) against severe Covid-19.

We observed lower vaccine effectiveness against hospitalization or death in individuals aged ≥75 years, compared to younger individuals, for a primary series of CoronaVac, a CoronaVac booster, and for a BNT162b2 booster (**Table 2**). However, vaccine effectiveness against hospitalization and death was significantly higher in individuals aged ≥75 years who received a heterologous BNT162b2 booster than a homologous CoronaVac booster ≥60 days of booster dose (78.5% vs 51.4%, respectively). Vaccine effectiveness against symptomatic disease was overall lower than severe COVID-19 across age groups and an age-related trend was not discernable.

**Table 2.** Effectiveness of homologous or heterologous booster against symptomatic Covid-19 and Covid-19 hospital admissions or deaths in adults stratified by age during Omicron period in Brazil

|  | <60 years |  | 60-74 years |  | ≥75 years |  |
| --- | --- | --- | --- | --- | --- | --- |
| **Symptomatic** | Controls/Cases | VE (95% CI) | Controls/Cases | VE (95% CI) | Controls/Cases | VE (95% CI) |
| Not vaccinated | 149451/172793 | Reference | 12394/17467 | Reference | 4005/6954 | Reference |
| Two doses, ≥180 days | 84833/102591 | 0.8% (-0.4-2) | 34007/39122 | 22% (19.8-24.2) | 15539/19463 | 28.1% (24.7-31.3) |
| Homologous booster |  |  |  |  |  |  |
| Third dose of CoronaVac, 8-59 days | 6058/6725 | 8.1% (4.6-11.4) | 1238/1493 | 12.2% (4.7-19) | 370/371 | 37.1% (26.7-46) |
| Third dose of CoronaVac, ≥60 days | 2578/3567 | -10.5% (-15.1--5.7) | 7352/9141 | 6.7% (2.7-10.5) | 11434/15139 | 13.2% (8.8-17.5) |
| Heterologous booster |  |  |  |  |  |  |
| Third dose of BNT162b2, 8-59 days | 108819/57507 | 57.3% (56.8-57.9) | 23479/15053 | 57.9% (56.5-59.3) | 4932/3273 | 62.5% (60.2-64.7) |
| Third dose of BNT162b2, ≥60 days | 183245/156962 | 31.8% (31.1-32.5) | 130148/116955 | 41.8% (40.3-43.3) | 64910/64495 | 45% (42.7-47.3) |
| **Hospitalization or Death** |  |  |  |  |  |  |
| Not vaccinated | 1169/2894 | Reference | 693/2108 | Reference | 877/2683 | Reference |
| Two doses, ≥180 days | 732/688 | 71% (65.9-75.3) | 2035/3180 | 63.4% (58.3-67.9) | 2750/5207 | 40.7% (33.7-46.9) |
| Homologous booster |  |  |  |  |  |  |
| Third dose of CoronaVac, 8-59 days | 34/16 | 86% (71.7-93.1) | 73/50 | 80.7% (68.6-88.1) | 49/51 | 47.7% (12.4-68.8) |
| Third dose of CoronaVac, ≥60 days | 23/21 | 82% (60.9-91.7) | 511/462 | 76.1% (70.8-80.4) | 2146/2964 | 51.4% (44.6-57.3) |
| Heterologous booster |  |  |  |  |  |  |
| Third dose of BNT162b2, 8-59 days | 775/183 | 92.1% (90.2-93.7) | 1203/537 | 88.4% (86.2-90.2) | 755/540 | 77.3% (73.1-80.8) |
| Third dose of BNT162b2, ≥60 days | 1397/370 | 90.2% (88.3-91.7) | 7441/3099 | 90.4% (89.1-91.5) | 9755/7063 | 78.5% (76.1-80.6) |

A total of 1,240,266 RT-PCR/antigen tests out of 1,308,364 eligible were selected into matched case-control pairs for the waning vaccine effectiveness in the Omicron period (**eTable 4**). Administration of a CoronaVac booster was associated with an increased VE against hospitalization or death relative to individuals who received their second dose ≥180 days previously (Table 3; rVE 8-59 days after third dose 47.1%, 95% CI 27.8 to 61.2), but minimal increase in VE against symptomatic disease (rVE 8-59 days after booster dose 4.9%, 95% CI 1.5 to 8.1). In addition, the additional protection gained by the booster dose against hospitalization or death waned after three months (rVE 90-119 days after booster dose 23.5%, 95% CI 12.4 to 33.1; rVE ≥120 days after booster dose 20.7%, 95% CI 10.1 to 30.0). In contrast, a BNT162b2 booster was associated with substantial increase in protection against hospitalization or death that was maintained for at least four months (**Table 3**; rVE ≥120 days after booster dose 62.8%, 95% CI 59.3 to 65.9). The gain in VE against symptomatic disease was lower for the homologous booster and appeared to wane over time for homologous and heterologous booster (**Table 3**).

**Table 3.**
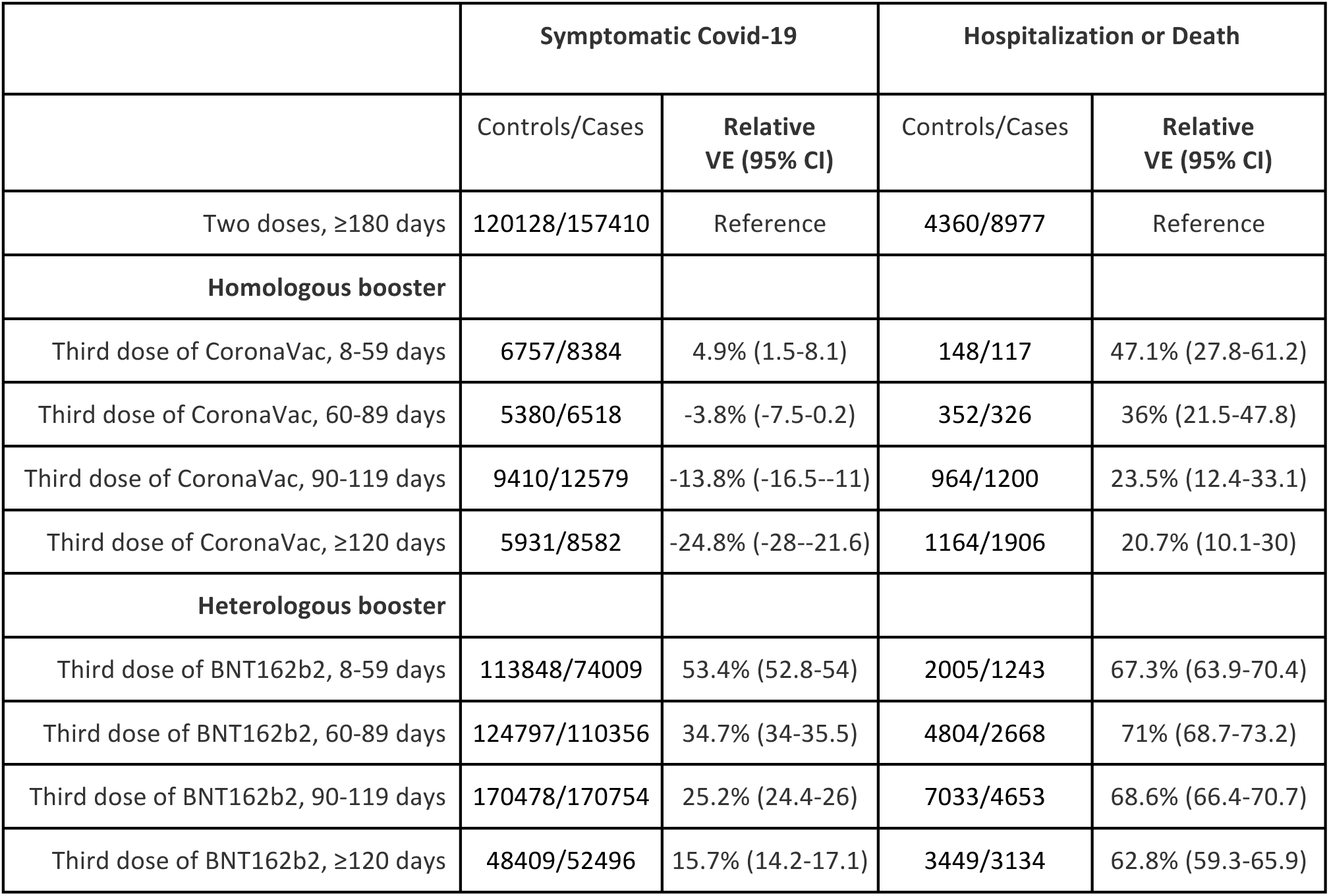
Vaccine effectiveness of a homologous and heterologous booster dose, relative to primary vaccination with CoronaVac during the period greater or equal to 180 days after the 2nd dose during Omicron period

|  | Symptomatic Covid-19 |  | Hospitalization or Death |  |
| --- | --- | --- | --- | --- |
|  | Controls/Cases | Relative VE (95% CI) | Controls/Cases | Relative VE (95% CI) |
| Two doses, $\geq 180$ days | 120128/157410 | Reference | 4360/8977 | Reference |
| **Homologous booster** |  |  |  |  |
| Third dose of CoronaVac, 8-59 days | 6757/8384 | 4.9% (1.5-8.1) | 148/117 | 47.1% (27.8-61.2) |
| Third dose of CoronaVac, 60-89 days | 5380/6518 | -3.8% (-7.5-0.2) | 352/326 | 36% (21.5-47.8) |
| Third dose of CoronaVac, 90-119 days | 9410/12579 | -13.8% (-16.5--11) | 964/1200 | 23.5% (12.4-33.1) |
| Third dose of CoronaVac, $\geq 120$ days | 5931/8582 | -24.8% (-28--21.6) | 1164/1906 | 20.7% (10.1-30) |
| **Heterologous booster** |  |  |  |  |
| Third dose of BNT162b2, 8-59 days | 113848/74009 | 53.4% (52.8-54) | 2005/1243 | 67.3% (63.9-70.4) |
| Third dose of BNT162b2, 60-89 days | 124797/110356 | 34.7% (34-35.5) | 4804/2668 | 71% (68.7-73.2) |
| Third dose of BNT162b2, 90-119 days | 170478/170754 | 25.2% (24.4-26) | 7033/4653 | 68.6% (66.4-70.7) |
| Third dose of BNT162b2, $\geq 120$ days | 48409/52496 | 15.7% (14.2-17.1) | 3449/3134 | 62.8% (59.3-65.9) |

A sensitivity analysis for the matching strategy obtained comparable estimates to the main analysis (**eTable 5**, **eTable 6**, and **eTable 7**), as after adjusting for month of the second dose (**eTable 8**), as a sensitivity analysis that was restricted to RT-PCR tests only (**eFigure 3**, **eTable 9**) obtained comparable estimates to the main analysis (**eTable 10**, **eTable 11**, **eTable 12** and **eTable 13**).

## Discussion

In this large observational study, we observed substantially lower effectiveness of a primary series of CoronaVac, and of a homologous CoronaVac and heterologous BNT162b2 booster dose, against symptomatic Covid-19 during an Omicron-dominated period compared to a Delta-dominated period. Effectiveness against severe outcomes was more similar between the two periods. In addition, a homologous booster dose conferred no additional protection against symptomatic disease during the Omicron-dominated period, and a moderate increase in protection against severe disease. Of note, the increased protection afforded by a homologous booster against severe disease does waned during the four month period after its administration. In contrast, the effectiveness of a heterologous BNT162b2 booster dose was substantially higher against symptomatic and severe disease, and protection against severe disease appeared to be durable up to four months.

Our findings have immediate implications for the current suggestion to administer homologous booster doses of inactivated vaccines in the context of the current global spread of the Omicron variant.^8^ There was overall a small benefit of a homologous booster and, for individuals aged ≥75 years, both the primary series and homologous booster afforded limited protection against severe disease (40-50%). However, a heterologous booster dose of BNT162b2 afforded a substantial increase in protection against severe disease in all age groups, including the elderly with age ≥75 years, compared to the primary series and some protection against symptomatic disease, albeit of uncertain duration. Although the direct comparison between homologous and heterologous booster is not straightforward because of the potential confounding factors between the individuals who received each vaccine (eTable 2), the main differences are region of residence and age, both factors accounted for in our adjustment. Additionally, based on the literature and magnitude of difference on the estimated VEs, it is unlikely the difference to be due to residual confounding. These findings suggest that the use of homologous CoronaVac as an option for booster doses,^8^ may need to be revisited, as preference to heterologous booster doses may be crucial to reducing morbidity and mortality associated with Omicron epidemics. Further research should investigate combinations of heterologous booster doses other than BNT162b2, including non-mRNA vaccines.

The reduced effectiveness of primary vaccination with CoronaVac and subsequent boosting schemes was observed primarily for symptomatic to moderate cases during the Omicron period. Low neutralizing antibody responses against the Omicron variant have been observed in individuals receiving two doses of CoronaVac^25–27^ and three doses of CoronaVac.^25,28^ A BNT162b2 booster dose has been shown to increase neutralizing antibodies against Omicron compared to a primary series of CoronaVac,^25,28^ and to a higher level than individuals who received a primary series of BNT162b2.^27^ The protection against severe disease for inactivated vaccines observed in this study speaks to the gaps in understanding of correlates of protection against severe disease, with a decoupling between measured neutralizing antibodies and clinical protection. This disparity has been observed for the primary series of CoronaVac, with moderate-to-high levels of protection against severe disease maintained beyond six months^5^ despite the lack of detectable neutralizing antibodies during this period.^29^

Our findings on the effectiveness against severe Covid-19 of homologous and heterologous booster doses during the Delta period is consistent with a previous test-negative study in Brazil^5^ and with a cohort study from Chile conducted during the same Delta period.^30^ For the Omicron period, our estimates are consistent with an ecological study from Hong Kong regarding the effectiveness of a primary vaccination with CoronaVac.^12^ However, our estimates of vaccine effectiveness against severe disease for a homologous booster are lower than reported the study in Hong Kong. The population seroprevalence in Brazil is higher than in Hong Kong, meaning that there is likely more infection-derived immunity in unvaccinated individuals, leading to lower VE estimates in this context. In addition, differences in study design, time of follow-up, non-pharmaceutical interventions in place during the Omicron outbreak in Hong Kong, and limited sample size for severe disease in the Hong Kong study could introduce differences.^12^

There was evidence for waning of effectiveness against symptomatic disease for homologous and heterologous boosters, and against severe disease for a homologous booster dose after three months during the Omicron period. This finding is consistent with numerous studies of primary series vaccination,^4–6^ and with more recent studies of booster dose effectiveness over time.^7,31^ In this study, we attempted to mitigate this bias by estimating relative VE over time since booster dose administration. We chose to evaluate waning using the reference group of those ≥180 days of second dose, assuming the waning from this period is slow or minimal. Where appreciable waning ≥180 days of second dose occurs, the interpretation of waning from rVE could be limited, because we would compare waning of booster against waning of second dose. Studies designed to identify and mitigate such biases should be prioritized to estimate the extent and timescale of waning effectiveness.^15,33^

We observed “negative” VE for some vaccination groups of homologous booster particularly for the Omicron period and homologous booster against symptomatic Covid-19. This phenomenon has been observed in some VE studies against Covid-19 and it is likely related to uncontrolled bias.^34^ We observed “negative” VE after some time from the vaccine, likely relating the bias driven by those early adopters or a widespread attack rate during Omicron surge. Additionally, some modeling suggests increased contact between vaccinated individuals associated with low VE could explain “negative” VE.^35^ Other unexpected finding is the lower VE in the groups <75 y compared to ≥75 y against symptomatic COVID-19. Differences in risk behaviour between age groups during the Omicron surge could explain these findings, making higher attack rates among the young and decreasing VE.^36^ This phenomenon was not observed for VE against severe Covid-19.

There are several strengths of our study. We used a nationwide database resulting in a large sample size and geographical coverage. We applied a matched test-negative design, including matching by time of epidemic and each one of 5,570 Brazilian municipalities. Finally, the timing of the booster campaign in Brazil together with the size and extent of the Omicron epidemic afforded us an opportunity to analyze a large population with three vaccine doses during an Omicron-dominated period, providing effectiveness estimates with relatively high precision even in age subgroups and over time.

Some limitations should be acknowledged. The data available for this study was collected as part of Brazil’s passive surveillance efforts for Covid-19, so important covariates may be missing or incomplete. As usually done in population-based studies with record linkage, we considered those individuals not linked to the vaccination database as unvaccinated, so we can have some degree of misclassification on vaccination status. We did not expect a relevant proportion of misclassification because the databases are centrally managed by the same data guarantor and we excluded few inconsistencies between and within databases. The distribution of RT-PCR tests and antigen tests, which have different sensitivity, changed over the course of the study period, which could have led to a decrease in estimated VE during the Omicron period through misclassification. However, a sensitivity analysis restricted to RT-PCR tests produced similar results. In the same topic, a higher vaccine effectiveness was observed in the main analysis against symptomatic Covid-19 during Omicron period (VE: 24.6%) in the period 0-13 days after the first dose (“bias indicator”) compared with the estimate n the sensitivity analysis restricted to RT-PCR tests (VE: 4.6%), showing evidence for potential misclassification of rapid antigen tests. In all analyses the overall vaccine effectiveness estimates were consistent and we don’t expect these biases would change considerably the message of this study. In addition, the test-negative controls may have been different during the Omicron and Delta periods, which could explain some of the difference in VE estimates. In particular, a higher proportion of controls were hospitalized or died during the Delta period (**Table 1**), implying either that other pathogens with severe outcomes were circulating during that period, or that less testing was being done in the outpatient setting during the Delta period. VE estimates in the primary analysis could be biased downwards due to accrual of undetected infection in unvaccinated individuals.^20,32^ However, if the additional protection by the vaccine on those with previous infection are proportional to the added protection in those naive of infection, the bias could be minimal.. We tried to adjust for it by using the proxy indication of previous COVID-19 likely illness, and at the same time immunity acquired by natural infection is less protective against Omicron. Our study is observational and so the VE is subject to confounding,^37^ given the bias indicator for the Delta period is close to 0, and for Omicron is significantly different from 0 for symptomatic cases, our VE estimates for the Omicron period appear to be more affected by bias.22 Additionally, the direct comparison between homologous and heterologous booster doses is not straightforward because there is a possibility for differences in risk by those who were offered and uptake of each booster type. However, we have controlled for important confounders (in particular age, location, time of test, comorbidities), and the difference in effectiveness observed between homologous and heterologous boosters is very unlikely to be explained by unmeasured confounding alone. Nevertheless, our results could be biased through this mechanism, and the direction of bias is unclear. Finally, differences in effectiveness and waning patterns by age could be driven by other factors, including occupational exposure (e.g., health care workers) and personal risk mitigation behavior.^15^

Overall, we found that primary vaccination with two doses of the CoronaVac vaccine provided 40-50% effectiveness against severe Covid-19 outcomes during the Omicron epidemic in Brazil, although effectiveness against symptomatic disease was close to zero. While a homologous booster afforded little additional protection, a heterologous booster dose of BNT162b2 restored high effectiveness against severe Covid-19, and moderate effectiveness against symptomatic disease up to four months.

## Contributors

All authors conceived the study. DATC, NED, JRA, AIK, and JC contributed equally as senior authors. OTR completed analyses with guidance from MDTH, MLL, DATC, NED, JRA, AIK, and JC. MSST, DHT, LCSD, MDTH and OTR curated and validated the data. OTR and MDTH wrote the first draft of the manuscript. RLM, GVAF, CFRF, MA, RFCS and RS provided supervision. All authors contributed to, and approved, the final manuscript. OTR and JC had verified the data. JC is the guarantor. The corresponding author attests that all listed authors meet authorship criteria and that no others meeting the criteria have been omitted.

## Data Availability

Data sharing: Deidentified databases as well as the R codes will be deposited in the repository https://github.com/juliocroda/VebraCOVID-19

## Acknowledgments

We thank the Pan American Health Organization for its support.

## Data availability

Deidentified databases will be deposited in the repository https://github.com/juliocroda/VebraCOVID-19

## Code availability

Code used to perform conduct statistical analysis will be available in the repository https://github.com/juliocroda/VebraCOVID-19

---

**eFigure 1.**
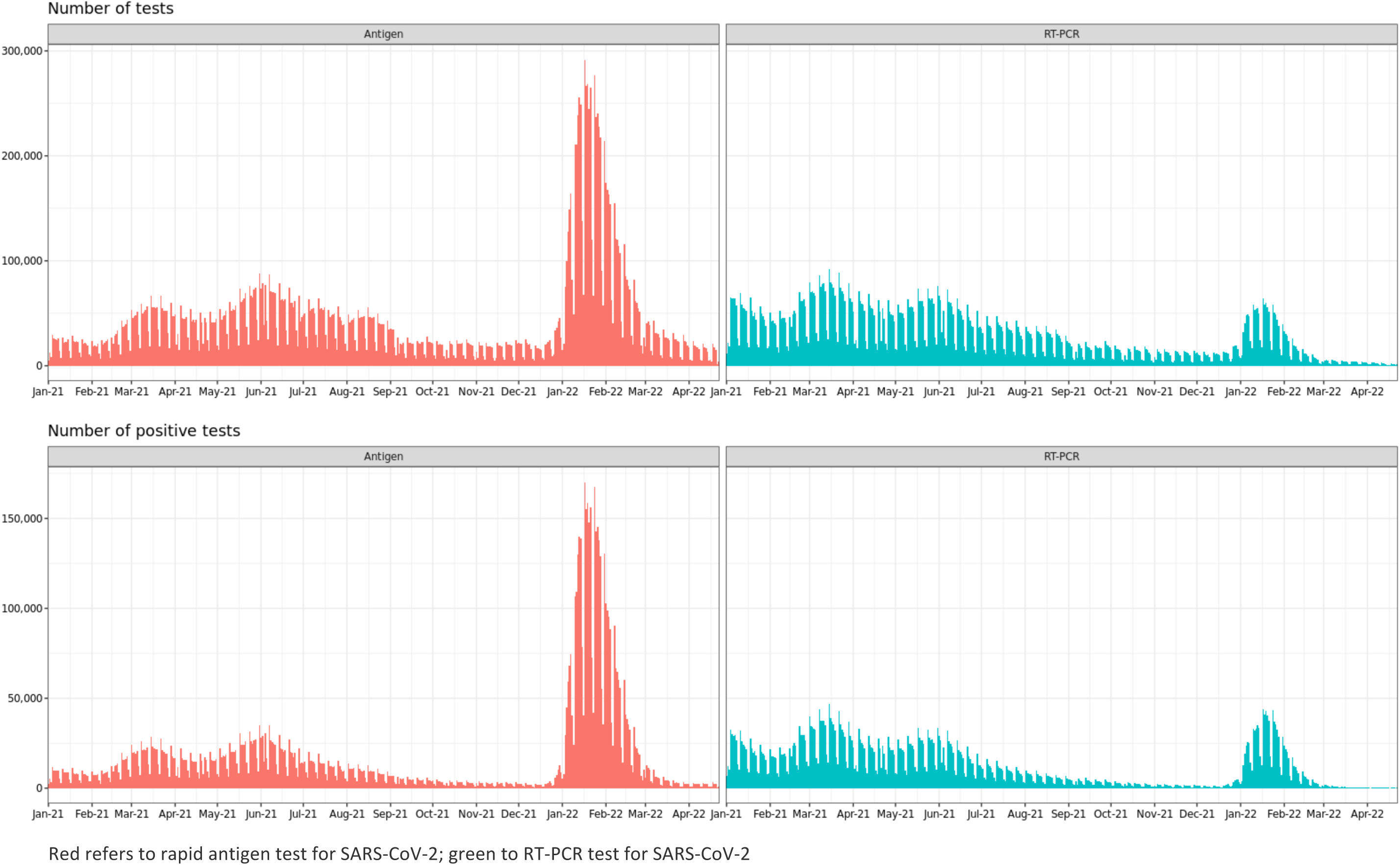
Number of SARS-CoV-2 RT-PCR and rapid antigen tests in symptomatic individuals performed in Brazil since January 2021.

**eFigure 2.**
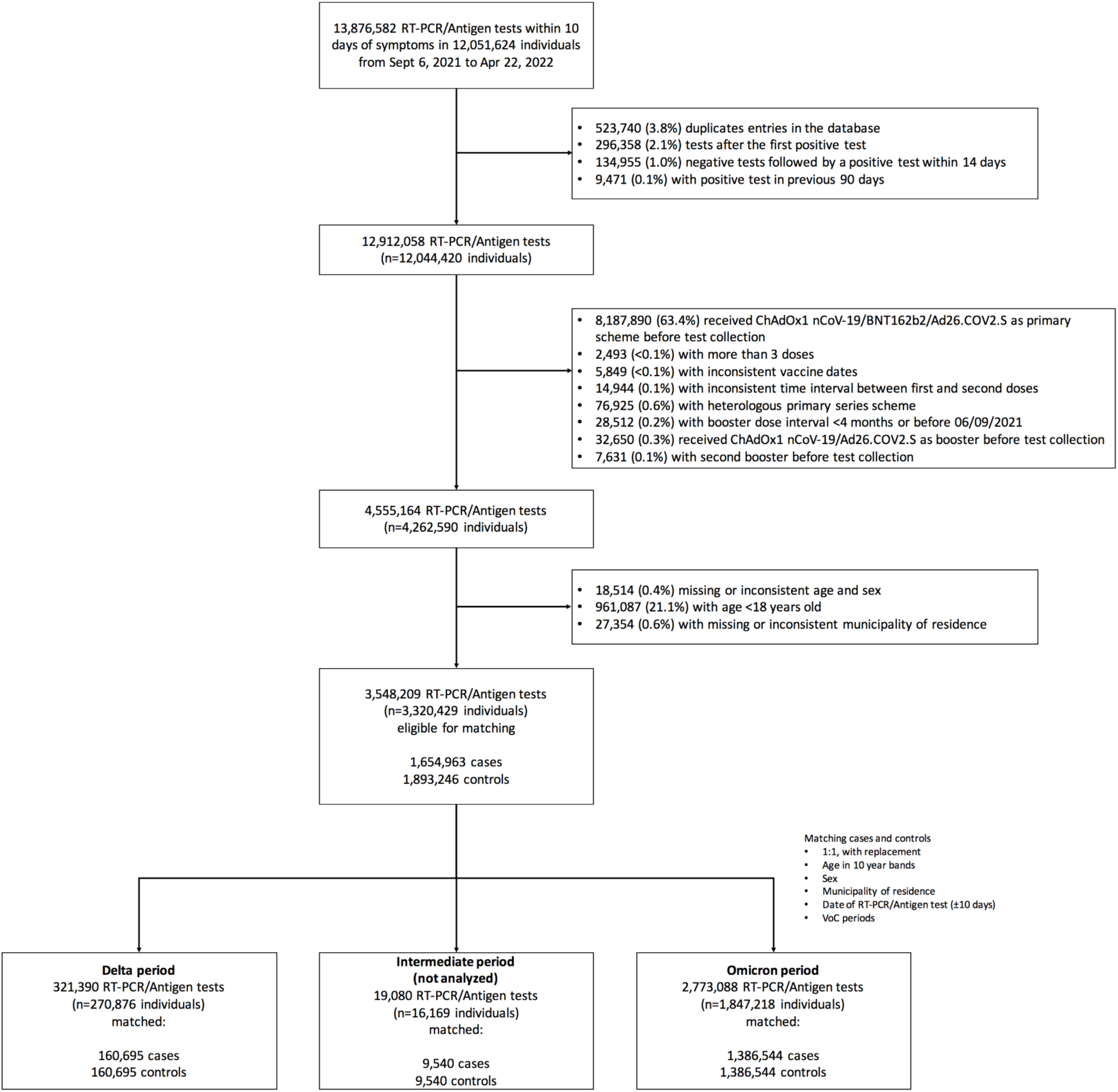
Study flow chart showing inclusion of cases and controls for the primary analysis

**eFigure 3.**
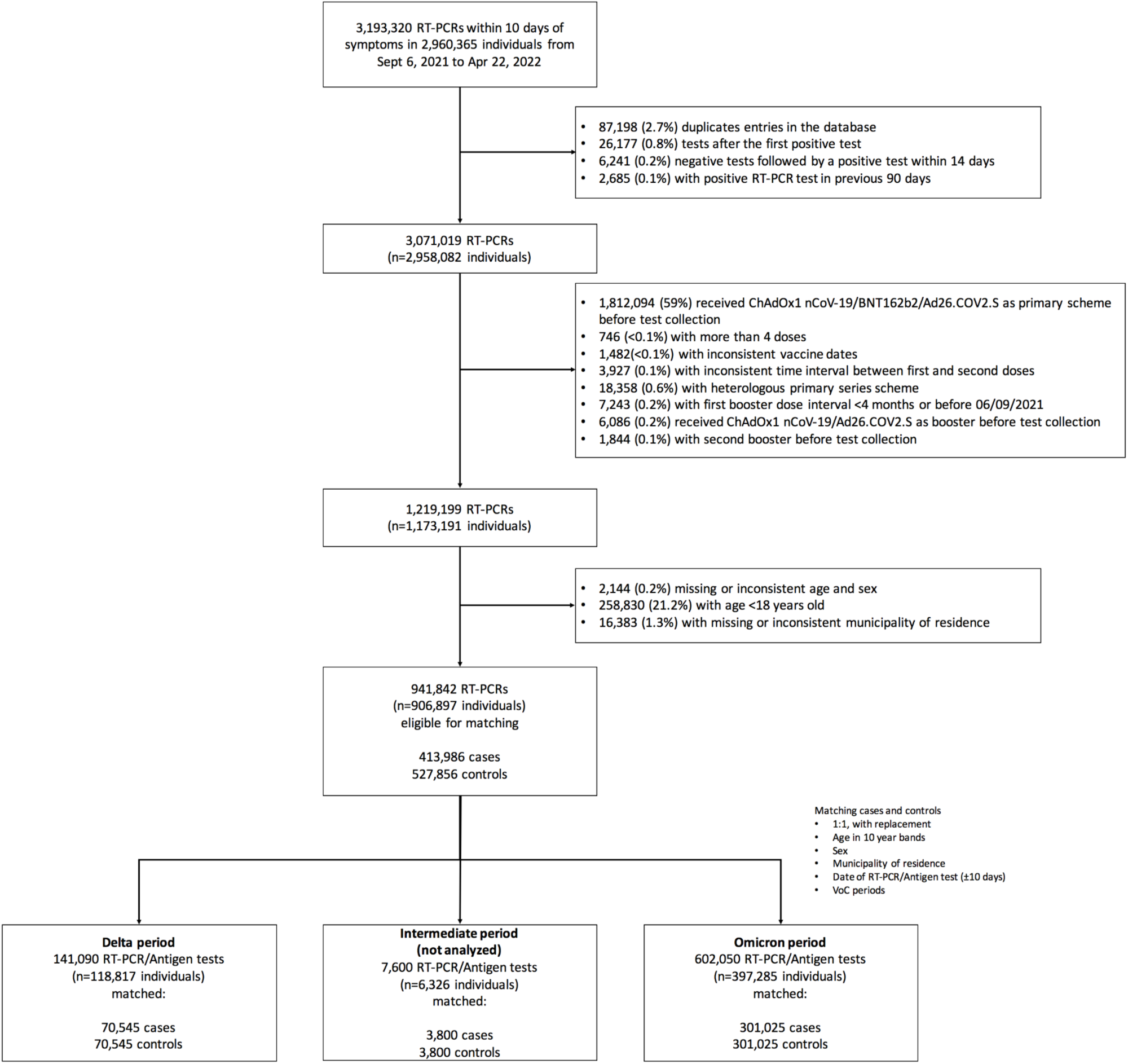
Flow chart showing inclusion of cases and controls for the sensitivity analysis including only RT-PCR SARS-CoV-2 tests

**eTable 1.** Data sources

| Data | Source | Extracted | Censored | Content |
| --- | --- | --- | --- | --- |
| Suspected mild COVID-19 | e-SUS notifica | 29/04/2022 | 22/04/2022 | COVID-19 suspected individuals of any age in Brazil, from public and private sectors, with mild clinical presentation, including those asymptomatic, not tested, those tested negative and tested positive. COVID-19 suspected cases notification is compulsory. |
| Suspected Severe COVID-19 | SIVEP-Gripe | 29/04/2022 | 22/04/2022 | Severe Acute Respiratory Infections (SARI) at any age in Brazil, from public and private sectors. Includes those not tested, those tested negative and those tested positive. It also includes SARI caused by other respiratory viruses, other aetiologies and those without an etiology. During COVID-19 pandemic, the official system to which COVID-19 hospital admissions and deaths must be notified. COVID-19 suspected cases notification is compulsory. |
| COVID-19 vaccination status | SI-PNI | 29/04/2022 | 22/04/2022 | National database from the Brazilian National Programme of Vaccination. COVID-19 vaccines notification is compulsory. |
| Variants of Concern in Brazil | GISAID | 09/05/2022 | 22/04/2022 | Public genomic repository data |

**eTable 2.** General characteristics of who received homologous or heterologous booster

|  | **Homologous booster**<br>**(CoronaVac)**<br>**(n=69,111)** | **Heterologous booster**<br>**(BNT152b2)**<br>**(n=984,444)** | **SMD** |
| --- | --- | --- | --- |
| **Demographics** |  |  |  |
| **Age, mean (SD), years** | 65.1 (21.5) | 53.2 (19.1) | 0.602 |
| **Age categories, n (%)** |  |  | 0.645 |
| **18-39 years** | 12890 (18.7) | 321694 (32.7) |  |
| **40-59 years** | 8313 (12.0) | 227987 (23.2) |  |
| **60-79 years** | 31925 (46.2) | 380315 (38.6) |  |
| **≥80 years** | 15983 (23.1) | 54448 (5.5) |  |
| **Male sex, n (%)** | 27431 (39.7) | 336163 (34.1) | 0.115 |
| **Self-reported race†, n (%),** |  |  | 0.399 |
| **White/Branca** | 42677 (61.8) | 461276 (46.9) |  |
| **Brown/Pardo** | 9464 (13.7) | 278390 (28.3) |  |
| **Black/Preta** | 1750 (2.5) | 34897 (3.5) |  |
| **Yellow/ Amarela** | 652 (0.9) | 15873 (1.6) |  |
| **Indigenous/Indigena** | 25 (0.0) | 2377 (0.2) |  |
| **Missing** | 14543 (21.0) | 191631 (19.5) |  |
| **Region of residence** |  |  | 1.234 |
| **North** | 278 (0.4) | 49932 (5.1) |  |
| **Northeast** | 302 (0.4) | 173982 (17.7) |  |
| **Central-West** | 350 (0.5) | 75923 (7.7) |  |
| **Southeast** | 66494 (96.2) | 489057 (49.7) |  |
| **South** | 1687 (2.4) | 195550 (19.9) |  |
| **Reported number of chronic comorbidities‡, n (%)** |  |  | 0.221 |
| **None** | 56094 (81.2) | 875843 (89.0) |  |
| **One or two** | 12457 (18.0) | 104856 (10.7) |  |
| **Three or more** | 560 (0.8) | 3745 (0.4) |  |
| **Prior SARS-CoV-2 exposure** |  |  |  |
| **Previous symptomatic events notified to the surveillance system¶, n (%)** | 14381 (20.8) | 253801 (25.8) | 0.118 |
| **Positive SARS-CoV-2 test result††, n (%)** | 3268 (4.7) | 52378 (5.3) | 0.027 |
| **Interval between symptoms onset and RT-PCR/Antigen testing, median (p25-p75), days** | 3 [2, 4] | 3.00 [2, 4] | 0.038 |
| **Interval between third dose and testing, median (p25-p75), days** | 93 [50, 116] | 87 [56, 105] | 0.123 |
| **Omicron period** | 67747 (98.0) | 966813 (98.2) | 0.013 |

RT-PCR=reverse transcription polymerase chain reaction; SMD=standardized mean difference; SD=standard deviation; SMD=standardized mean difference. † Race/skin colour as defined by the Brazilian national census bureau (Instituto Nacional de Geografia e Estatísticas). ‡ Comorbidities included cardiovascular, or renal conditions, diabetes, chronic respiratory disorder, obesity, or immunosuppression. ¶ Reported illness with covid-19 associated symptoms in eSUS and SIVEP-Gripe databases before the start of study on 06 September 2021.. †† Defined as a positive SARS-CoV-2 RT-PCR or antigen detection test result before the start of study on 06 September 2021.

**eTable 3.** Effectiveness of CoronaVac and homologous or heterologous booster against symptomatic Covid-19 and hospital admissions or deaths in adults in Brazil

|  | Symptomatic COVID-19 |  | Hospitalization or death |  |
| --- | --- | --- | --- | --- |
|  | Delta | Omicron | Delta | Omicron |
| **Not vaccinated** | Reference | Reference | Reference | Reference |
| **Single dose, within 0-13 days** | 2.2% (-4-8.1) | 24.6% (19.1-29.8) | 1.7% (-28.4-30.8) | -3.1% (-57.5-54.7) |
| **Single dose, ≥14 days** | 27.2% (25.1-29.3) | 17% (15.9-18) | 46.6% (38.9-53.3) | 47.5% (40.7-53.5) |
| **Two doses, within 0-13 days** | 38.5% (35.3-41.5) | 21.3% (17.3-25.2) | 76.7% (67.3-83.4) | 14.8% (-46-60.8) |
| **Two doses, 14-59 days** | 51.3% (49.9-52.7) | 28.1% (26.5-29.6) | 86.5% (83.4-88.9) | 56.1% (40.6-67.5) |
| **Two doses, 60-179 days** | 37.6% (36.1-39.1) | 3.9% (3.1-4.8) | 71% (68.5-73.2) | 61.4% (57.4-65) |
| **Two doses, ≥180 days** | 34% (32.3-35.7) | 6.3% (5.3-7.3) | 60.9% (57.3-64.2) | 57.6% (54.4-60.6) |
| **Third dose of CoronaVac, 0-7 days** | 44.7% (26.2-58.5) | 9.6% (1.7-16.9) | 80.8% (67.7-88.6) | 75.6% (26.7-91.8) |
| **Third dose of CoronaVac, 8-59 days** | 57.1% (50.4-62.9) | 8.6% (5.6-11.5) | 75.9% (67.8-81.9) | 73.6% (63.9-80.7) |
| **Third dose of CoronaVac, ≥60 days** | 53.5% (30.9-68.6) | -2.9% (-5.2--0.6) | 75.6% (52.8-87.4) | 67.8% (64.3-71) |
| **Third dose of BNT162b2, 0-7 days** | 42.6% (38.5-46.5) | 16.6% (14.7-18.4) | 80.4% (75.8-84) | 62.6% (50.7-71.6) |
| **Third dose of BNT162b2, 8-59 days** | 86.7% (86-87.4) | 56.8% (56.3-57.3) | 92.3% (91-93.4) | 86% (84.5-87.4) |
| **Third dose of BNT162b2, ≥60 days** | 83.2% (79.8-86.1) | 33.8% (33.2-34.4) | 90.8% (84.8-94.4) | 86.4% (85.4-87.3) |

**eTable 4.** Characteristics of adults in Brazil, who were selected into case test negative pairs for the CoronaVac analysis during the Delta period (September 6, 2021 to December 14, 2021) and Omicron period (December 25, 2021 to April 22, 2022), for the analysis of relative vaccine effectiveness

|  | Matched pairs for Omicron period |  |  |
| --- | --- | --- | --- |
|  | Controls (n=620,133) | Cases (n=620,133) | SMD |
| **Demographics** |  |  |  |
| **Age, mean (SD), years** | 54.23 (18.9) | 54.5 (19.1) | 0.016 |
| **Age categories, n (%)** |  |  | 0.058 |
| 18-39 years | 185374 (29.9) | 181268 (29.2) |  |
| 40-59 years | 150442 (24.3) | 154755 (25.0) |  |
| 60-79 years | 247298 (39.9) | 238998 (38.5) |  |
| ≥80 years | 37019 ( 6.0) | 45112 ( 7.3) |  |
| **Male sex, n (%)** | 214071 (34.5) | 214071 (34.5) | <0.001 |
| **Self-reported race<sup>†</sup>, n (%)** |  |  | 0.052 |
| White/Branca | 289223 (46.6) | 293911 (47.4) |  |
| Brown/Pardo | 178933 (28.9) | 168763 (27.2) |  |
| Black/Preta | 23067 ( 3.7) | 21092 ( 3.4) |  |
| Yellow/ Amarela | 9970 ( 1.6) | 9487 ( 1.5) |  |
| Indigenous/Indigena | 2818 ( 0.5) | 2098 ( 0.3) |  |
| Missing | 116122 (18.7) | 124782 (20.1) |  |
| **Region of residence** |  |  | <0.001 |
| North | 33438 ( 5.4) | 33438 ( 5.4) |  |
| Northeast | 100347 (16.2) | 100347 (16.2) |  |
| Central-West | 45755 ( 7.4) | 45755 ( 7.4) |  |
| Southeast | 328824 (53.0) | 328824 (53.0) |  |
| South | 111769 (18.0) | 111769 (18.0) |  |
| **Reported number of chronic comorbidities<sup>‡</sup>, n (%)** |  |  | 0.014 |
| None | 546266 (88.1) | 548356 (88.4) |  |
| One or two | 71314 (11.5) | 68945 (11.1) |  |
| Three or more | 2553 ( 0.4) | 2832 ( 0.5) |  |
| **Prior SARS-CoV-2 exposure** |  |  |  |
| Previous symptomatic events notified to the surveillance system<sup>¶</sup>, n (%) | 164774 (26.6) | 151882 (24.5) | 0.124 |
| Positive SARS-CoV-2 test result<sup>††</sup>, n (%) | 40970 ( 6.6) | 23827 ( 3.8) | 0.127 |
| **Interval between symptoms onset and RT-PCR testing, median (p25-p75), days** | 3 [2, 4] | 3 [2, 4] | 0.127 |
| **Hospitalization or Death** | 15771 ( 2.5) | 24395 ( 3.9) | 0.079 |
| **Vaccination status** |  |  | 0.223 |
| Two doses, ≥180 days, n (%) | 120128 (19.4) | 157410 (25.4) |  |
| Third dose of CoronaVac, 0-7 days, n (%) | 878 ( 0.1) | 1140 ( 0.2) |  |
| Third dose of CoronaVac, 8-59 days, n (%) | 6757 ( 1.1) | 8384 ( 1.4) |  |
| Third dose of CoronaVac, 60-89 days, n (%) | 5380 ( 0.9) | 6518 ( 1.1) |  |
| Third dose of CoronaVac, 90-119 days, n (%) | 9410 (1.5) | 12579 ( 2.0) |  |
| Third dose of CoronaVac, ≥90 days, n (%) | 5931 (1.0) | 8582 ( 1.4) |  |
| Third dose of BNT162b2, 0-7 days, n (%) | 14117 ( 2.3) | 17905 ( 2.9) |  |
| Third dose of BNT162b2, 8-59 days, n (%) | 113848 (18.4) | 74009 (11.9) |  |
| Third dose of BNT162b2, 60-89 days, n (%) | 124797 (20.1) | 110356 ( 17.8) |  |
| Third dose of BNT162b2, 90-119 days, n (%) | 170478 (27.5) | 170754 ( 27.5) |  |
| Third dose of BNT162b2, ≥120 days, n (%) | 48409 (7.8) | 52496 ( 8.5) |  |
| **Interval between second dose and RT-PCR/Antigen test, mean (SD), days** | 269 [228, 305] | 269 [223, 306] | 0.012 |
| **Interval between third dose and RT-PCR/Antigen test, mean (SD), days** | 87 [56, 105] | 91 [65, 109] | 0.133 |

RT-PCR=reverse transcription polymerase chain reaction; SMD=standardized mean difference; SD=standard deviation; † Race/skin colour as defined
by the Brazilian national census bureau (Instituto Nacional de Geografia e Estatísticas). ‡ Comorbidities included cardiovascular, or renal conditions, diabetes, chronic respiratory disorder, obesity, or immunosuppression. ¶ Reported illness with covid-19 associated symptoms in eSUS and SIVEP-
Gripe databases before the start of study on 06 September 2021. †† Defined as a positive SARS-CoV-2 RT-PCR or antigen detection test result before the start of study on 06 September 2021..

**eTable 5.** Characteristics of adults in Brazil, who were selected into case test negative pairs for the analysis of vaccine effectiveness during the Delta period (September 6, 2021 to December 14, 2021) and the Omicron period (December 25, 2021 to April 22, 2022) - Sensitivity analysis for matching

|  | Matched pairs for Delta period |  |  | Matched pairs for Omicron period |  |  |
| --- | --- | --- | --- | --- | --- | --- |
|  | Controls<br>(n=360,045) | Cases<br>(n=132,355) | SMD | Controls<br>(n=982,144) | Cases<br>(n=1,213,302) | SMD |
| **Demographics** |  |  |  |  |  |  |
| **Age, mean (SD), years** | 40.4 (17.9) | 44.6 (19.4) | 0.225 | 41.7 (18.5) | 42.2 (18.4) | 0.025 |
| **Age categories, n (%)** |  |  | 0.227 |  |  | 0.034 |
| 18-39 years | 222864 (61.9) | 68163 (51.5) |  | 573177 (58.4) | 696053 (57.4) |  |
| 40-59 years | 68145 (18.9) | 27948 (21.1) |  | 196976 (20.1) | 256255 (21.1) |  |
| 60-79 years | 61653 (17.1) | 31947 (24.1) |  | 187718 (19.1) | 227151 (18.7)) |  |
| ≥80 years | 7383 ( 2.1) | 4297 ( 3.2) |  | 24273 ( 2.5) | 33843 ( 2.8) |  |
| **Male sex, n (%)** | 148640 (41.3) | 59822 (45.2) | 0.079 | 388595 (39.6) | 495040 (40.8) | 0.025 |
| **Self-reported race<sup>†</sup>, n (%)** |  |  | 0.071 |  |  | 0.087 |
| White/Branca | 165235 (45.9) | 58768 (44.4) |  | 431055 (43.9) | 540505 (44.5) |  |
| Brown/Pardo | 99280 (27.6) | 37519 (28.3) |  | 306878 (31.2) | 346765 (28.6) |  |
| Black/Preta | 15917 ( 4.4) | 5106 ( 3.9) |  | 43312 ( 4.4) | 46625 ( 3.8) |  |
| Yellow/ Amarela | 4090 ( 1.1) | 1861 ( 1.4) |  | 17671 ( 1.8) | 20763 ( 1.7) |  |
| Indigenous/Indigena | 1101 ( 0.3) | 898 ( 0.7) |  | 2800 ( 0.3) | 2233 ( 0.2) |  |
| Missing | 74422 (20.7) | 28203 (21.3) |  | 180428 (18.4) | 256411 (21.1) |  |
| **Region of residence** |  |  | 0.216 |  |  | 0.031 |
| North | 14887 ( 4.1) | 8488 ( 6.4) |  | 40809 ( 4.2) | 55841 ( 4.6) |  |
| Northeast | 34766 ( 9.7) | 15319 (11.6) |  | 135724 (13.8) | 168228 (13.9) |  |
| Central-West | 20801 ( 5.8) | 11562 ( 8.7) |  | 78255 ( 8.0) | 89169 ( 7.3) |  |
| Southeast | 211159 (58.6) | 64695 (48.9) |  | 541177 (55.1) | 668797 (55.1) |  |
| South | 78432 (21.8) | 32291 (24.4) |  | 186179 (19.0) | 231267 (19.1) |  |
| **Reported number of chronic comorbidities<sup>‡</sup>, n (%)** |  |  | 0.122 |  |  | 0.014 |
| None | 325084 (90.3) | 114438 (86.5) |  | 908535 (92.5) | 1126250 (92.8) |  |
| One or two | 33518 ( 9.3) | 16900 (12.8) |  | 71502 ( 7.3) | 84147 (6.9) |  |
| Three or more | 1443 ( 0.4) | 1017 ( 0.8) |  | 2107(0.2) | 2905 (0.2) |  |
| **Prior SARS-CoV-2 exposure** |  |  |  |  |  |  |
| Previous symptomatic events notified to the surveillance system<sup>¶</sup>, n (%) | 117610 (32.7) | 25033 (18.9) | 0.318 | 307779 (31.3) | 310788 (25.6) | 0.127 |
| Positive SARS-CoV-2 test result<sup>††</sup>, n (%) | 24952 ( 6.9) | 2212 ( 1.7) | 0.261 | 79126 ( 8.1) | 58724 ( 4.8) | 0.131 |
| **Interval between symptoms onset and RT-PCR testing, median (p25-p75), days** | 3 [2, 4] | 3 [2, 5] | 0.143 | 3 [2, 4] | 3 [2, 4] | 0.105 |
| **Hospitalization or death** | 9980 ( 2.8) | 14547 (11.0) | 0.329 | 13538 ( 1.4) | 28387 ( 2.3) | 0.071 |
| **Vaccination status** |  |  | 0.422 |  |  | 0.217 |
| Not vaccinated, n (%) | 73296 (20.4) | 40165 (30.3) |  | 115379 (11.7) | 168224 (13.9) |  |
| *Primary vaccination with CoronaVac* |  |  |  |  |  |  |
| Single dose, 0-13 days, n (%) | 4477 ( 1.2) | 2135 ( 1.6) |  | 1025 ( 0.1) | 1298 ( 0.1) |  |
| Single dose, ≥14 days, n (%) | 32487 ( 9.0) | 11580 ( 8.7) |  | 53933 ( 5.5) | 64038 ( 5.3) |  |
| Two doses, 0-13 days, n (%) | 11889 ( 3.3) | 3205 ( 2.4) |  | 2803 (0.2) | 2657(0.2) |  |
| Two doses, 14-89 days, n (%) | 92035 (25.6) | 21210 (16.0) |  | 41152 ( 4.2) | 44794 ( 3.7) |  |
| Two doses, 90-179 days, n (%) | 65840 (18.3) | 29431 (22.2) |  | 268947 (27.4) | 391160 (32.2) |  |
| Two doses, ≥180 days, n (%) | 47556 (13.2) | 20348 (15.4) |  | 98996 (10.1) | 137720 (11.4) |  |
| *Booster vaccination* |  |  |  |  |  |  |
| Third dose of CoronaVac, 0-7 days, n (%) | 225 ( 0.1) | 81 ( 0.1) |  | 647 ( 0.1) | 1072 ( 0.1) |  |
| Third dose of CoronaVac, 8-59 days, n (%) | 1384 ( 0.4) | 349 ( 0.3) |  | 5202 ( 0.5) | 7414 ( 0.6) |  |
| Third dose of CoronaVac, ≥60 days, n (%) | 241 ( 0.1) | 61 ( 0.0) |  | 12747 ( 1.3) | 21818 ( 1.8) |  |
| Third dose of BNT162b2, 0-7 days, n (%) | 4041 ( 1.1) | 1573 ( 1.2) |  | 11747 ( 1.2) | 16677 ( 1.4) |  |
| Third dose of BNT162b2, 8-59 days, n (%) | 24942 ( 6.9) | 2046 ( 1.5) |  | 96675 ( 9.8) | 66339 ( 5.5) |  |
| Third dose of BNT162b2, ≥60 days, n (%) | 1632 ( 0.5) | 171 ( 0.1) |  | 273307 (27.8) | 290091 (23.9) |  |
| **Interval between first dose and testing, median (p25-p75), days** | 34 [21, 62] | 33 [19, 62] | 0.032 | 138 [95, 169] | 140 [99, 170] | 0.033 |
| **Interval between second dose and testing, median (p25-p75), days** | 96 [45, 172] | 145 [66, 184] | 0.283 | 138 [115, 177] | 141 [119, 176] | 0.048 |
| **Interval between third dose and testing, median (p25-p75), days** | 24 [13, 41] | 10 [6, 29] | 0.451 | 86 [53, 107] | 91 [64, 107] | 0.073 |

RT-PCR=reverse transcription polymerase chain reaction; SMD=standardized mean difference; SD=standard deviation; † Race/skin colour as defined by the Brazilian national census bureau (Instituto Nacional de Geografia e
Estatísticas). ‡ Comorbidities included cardiovascular, or renal conditions, diabetes, chronic respiratory disorder, obesity, or immunosuppression. . ¶ Reported illness with covid-19 associated symptoms in eSUS and SIVEP-Gripe
databases before the start of study on 06 September 2021. \*\* Defined as a positive SARS-CoV-2 RT-PCR or antigen detection test result before the start of study on 06 September 2021

**eTable 6.** Sensitivity analysis of matching strategy to evaluate the vaccine effectiveness of CoronaVac and homologous or heterologous booster against symptomatic Covid-19 using RT-PCR or Antigen tests

|  | Delta Period |  | Omicron Period |  |
| --- | --- | --- | --- | --- |
|  | Matching 1:1 | Stratified by sets | Matching 1:1 | Stratified by sets |
| Not vaccinated | Reference | Reference | Reference | Reference |
| Single dose, within 0-13 days | 2.2% (-4-8.1) | 4.6% (-1.3-10.2) | 24.6% (19.1-29.8) | 17.8% (10.1-24.9) |
| Single dose, ≥14 days | 27.2% (25.1-29.3) | 23.9% (21.8-26) | 17% (15.9-18) | 14.6% (13.3-15.9) |
| Two doses, within 0-13 days | 38.5% (35.3-41.5) | 35.9% (32.8-38.8) | 21.3% (17.3-25.2) | 27.2% (22.6-31.4) |
| Two doses, 14-59 days | 51.3% (49.9-52.7) | 48.3% (46.9-49.7) | 28.1% (26.5-29.6) | 26.5% (24.6-28.4) |
| Two doses, 60-179 days | 37.6% (36.1-39.1) | 33.2% (31.7-34.7) | 3.9% (3.1-4.8) | 0.6% (-0.4-1.6) |
| Two doses, ≥180 days | 34% (32.3-35.7) | 29.7% (27.9-31.5) | 6.3% (5.3-7.3) | 3.6% (2.4-4.9) |
| Third dose of CoronaVac, 0-7 days | 44.7% (26.2-58.5) | 50.4% (34.2-62.7) | 9.6% (1.7-16.9) | -0.5% (-10.5-9.6) |
| Third dose of CoronaVac, 8-59 days | 57.1% (50.4-62.9) | 63.6% (58-68.5) | 8.6% (5.6-11.5) | 9.8% (6-13.3) |
| Third dose of CoronaVac, ≥60 days | 53.5% (30.9-68.6) | 60.1% (42.7-72.2) | -2.9% (-5.2--0.6) | -5.8% (-8.6--2.9) |
| Third dose of BNT162b2, 0-7 days | 42.6% (38.5-46.5) | 36.1% (31.6-40.3) | 16.6% (14.7-18.4) | 13.9% (11.5-16.1) |
| Third dose of BNT162b2, 8-59 days | 86.7% (86-87.4) | 86.4% (85.6-87.1) | 56.8% (56.3-57.3) | 55.2% (54.6-55.9) |
| Third dose of BNT162b2, ≥60 days | 83.2% (79.8-86.1) | 81.1% (77.2-84.3) | 33.8% (33.2-34.4) | 29.9% (29.1-30.7) |

**eTable 7.**
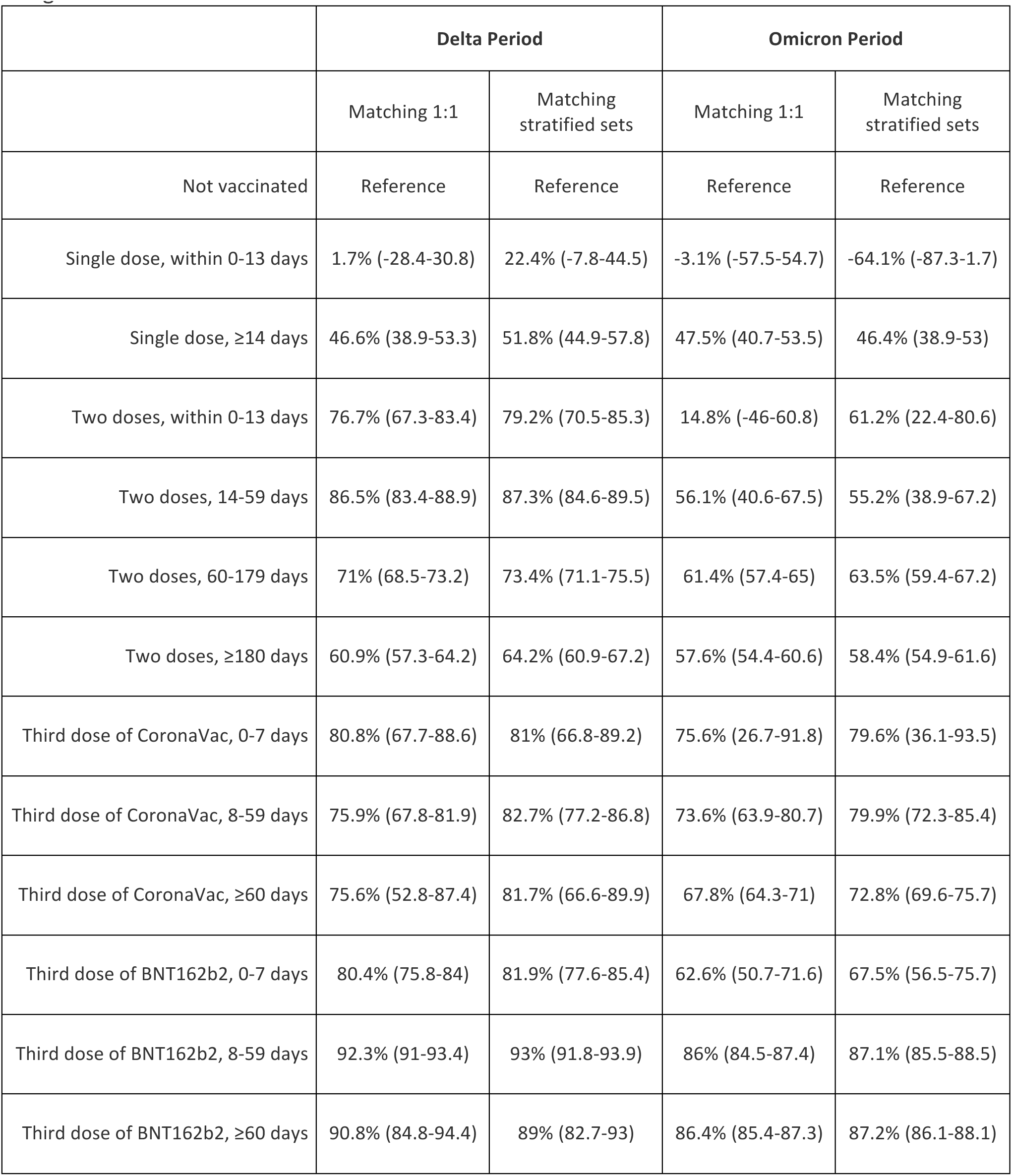
Sensitivity analysis of matching strategy to evaluate the vaccine effectiveness of CoronaVac and homologous or heterologous booster against Severe Covid-19 using RT-PCR or Antigen tests

**eTable 8.** Vaccine effectiveness of a homologous and heterologous booster dose, relative to primary vaccination with CoronaVac during the period greater or equal to 180 days after the 2nd dose during Omicron period, further adjusted by month of second dose (sensitivity analysis)

|  | Symptomatic Covid-19 |  | Hospitalization or Death |  |
| --- | --- | --- | --- | --- |
|  | Main analysis | Sensitivity analysis adjusted by month of 2nd dose | Main analysis | Sensitivity analysis adjusted by month of 2nd dose |
|  | Relative VE (95% CI) | Relative VE (95% CI) | Relative VE (95% CI) | Relative VE (95% CI) |
| **Two doses, ≥180 days** | Reference | Reference | Reference | Reference |
| **Homologous booster** |  |  |  |  |
| **Third dose of CoronaVac, 8-59 days** | 4.9% (1.5-8.1) | 1.4% (-2.1-4.8) | 47.1% (27.8-61.2) | 45.8% (26-60.3) |
| **Third dose of CoronaVac, 60-89 days** | -3.8% (-7.5-0.2) | -4.5% (-8.3--0.6) | 36% (21.5-47.8) | 35.8% (21.3-47.7) |
| **Third dose of CoronaVac, 90-119 days** | -13.8% (-16.5--11) | -14% (-16.7--11.1) | 23.5% (12.4-33.1) | 25.1% (14.3-34.6) |
| **Third dose of CoronaVac, ≥120 days** | -24.8% (-28--21.6) | -23.4% (-26.6--20.1) | 20.7% (10.1-30) | 24.3% (14-33.4) |
| **Heterologous booster** |  |  |  |  |
| **Third dose of BNT162b2, 8-59 days** | 53.4% (52.8-54) | 51.7% (51.1-52.4) | 67.3% (63.9-70.4) | 66.7% (63.2-69.9) |
| **Third dose of BNT162b2, 60-89 days** | 34.7% (34-35.5) | 34.5% (33.7-35.3) | 71% (68.7-73.2) | 70.9% (68.5-73) |
| **Third dose of BNT162b2, 90-119 days** | 25.2% (24.4-26) | 25% (24.1-25.9) | 68.6% (66.4-70.7) | 69.1% (66.8-71.1) |
| **Third dose of BNT162b2, ≥120 days** | 15.7% (14.2-17.1) | 16.9% (15.4-18.5) | 62.8% (59.3-65.9) | 64.3% (60.9-67.5) |

**eTable 9.**
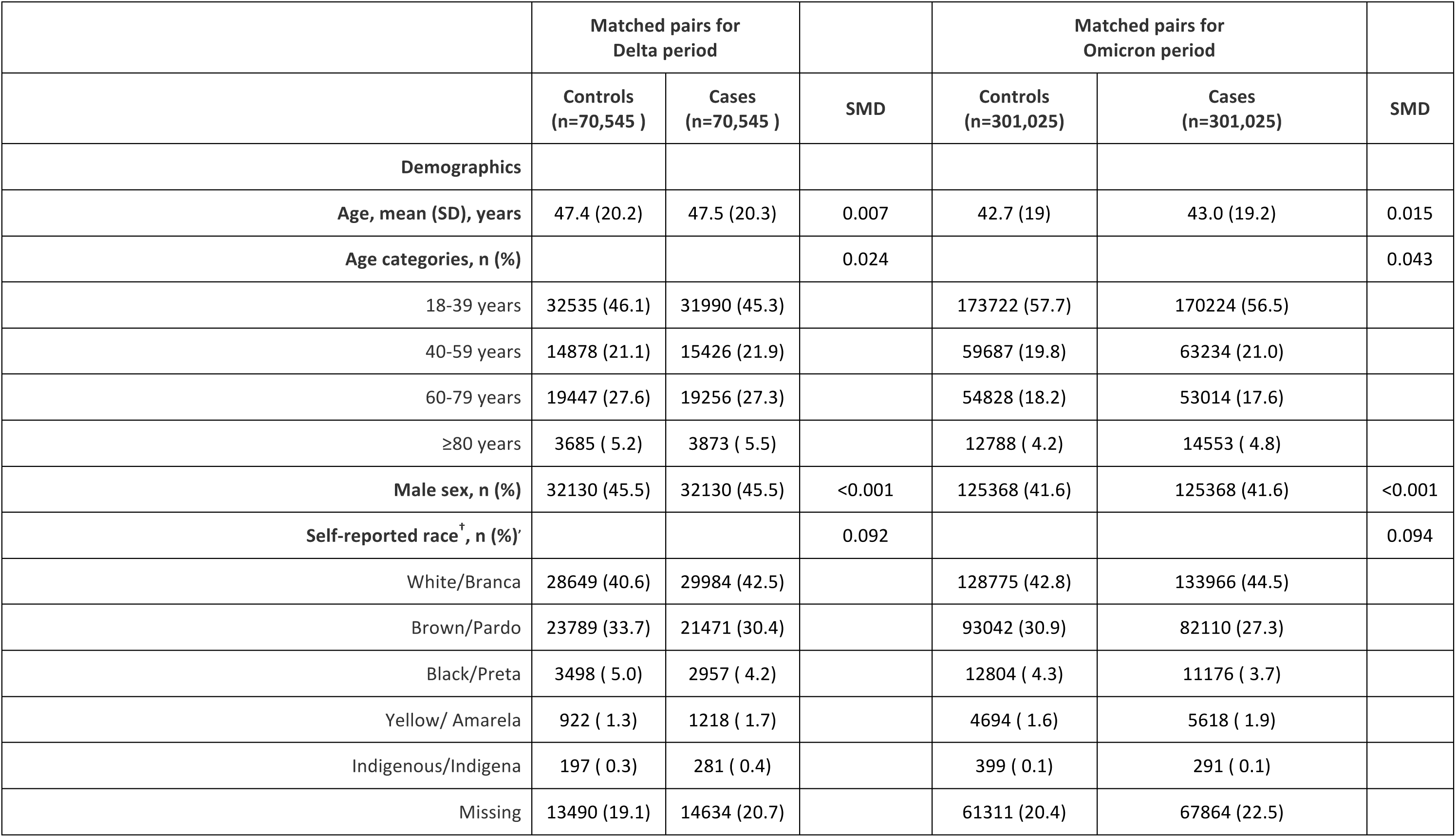
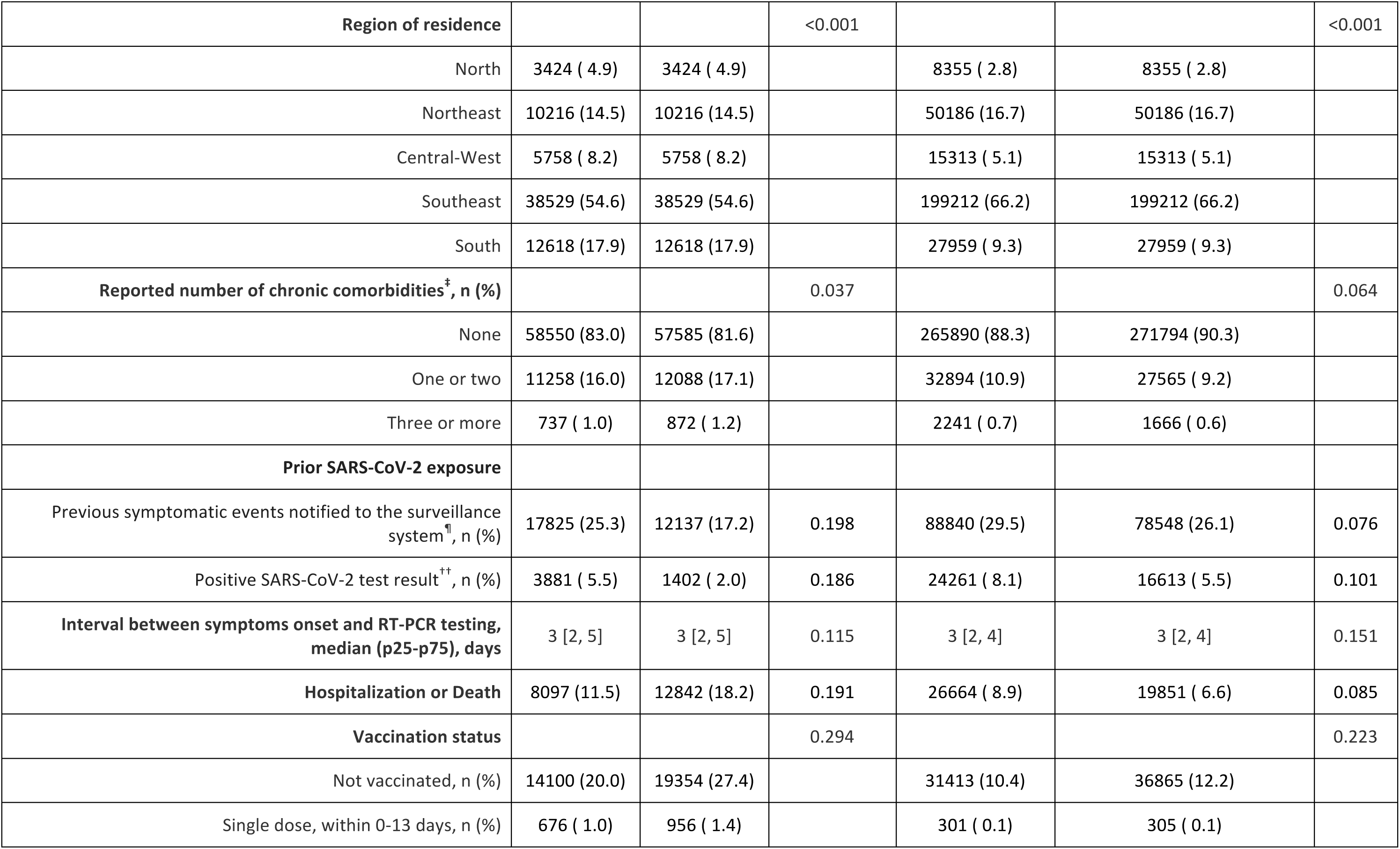
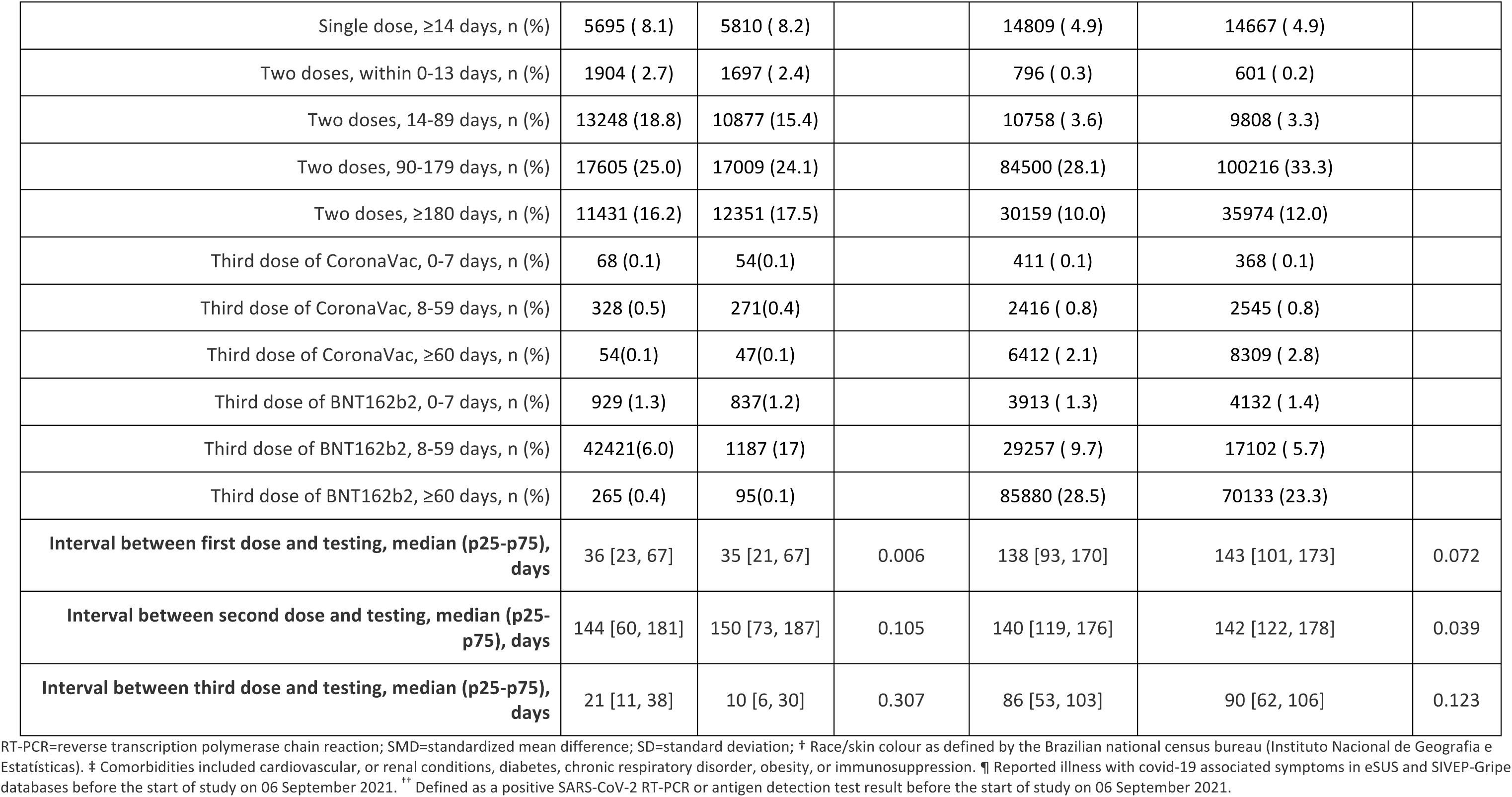
Characteristics of adults in Brazil, who were selected into case-test negative pairs for the sensitivity analysis including only RT-PCR tests during the Delta period (September 6, 2021 to December 14, 2021) and Omicron period (December 25, 2021 to April 2, 2022)

|  | Matched pairs for<br>Delta period |  |  | Matched pairs for<br>Omicron period |  |  |
| --- | --- | --- | --- | --- | --- | --- |
|  | Controls<br>(n=70,545 ) | Cases<br>(n=70,545 ) | SMD | Controls<br>(n=301,025) | Cases<br>(n=301,025) | SMD |
| **Demographics** |  |  |  |  |  |  |
| **Age, mean (SD), years** | 47.4 (20.2) | 47.5 (20.3) | 0.007 | 42.7 (19) | 43.0 (19.2) | 0.015 |
| **Age categories, n (%)** |  |  | 0.024 |  |  | 0.043 |
| 18-39 years | 32535 (46.1) | 31990 (45.3) |  | 173722 (57.7) | 170224 (56.5) |  |
| 40-59 years | 14878 (21.1) | 15426 (21.9) |  | 59687 (19.8) | 63234 (21.0) |  |
| 60-79 years | 19447 (27.6) | 19256 (27.3) |  | 54828 (18.2) | 53014 (17.6) |  |
| ≥80 years | 3685 ( 5.2) | 3873 ( 5.5) |  | 12788 ( 4.2) | 14553 ( 4.8) |  |
| **Male sex, n (%)** | 32130 (45.5) | 32130 (45.5) | <0.001 | 125368 (41.6) | 125368 (41.6) | <0.001 |
| **Self-reported race<sup>†</sup>, n (%)** |  |  | 0.092 |  |  | 0.094 |
| White/Branca | 28649 (40.6) | 29984 (42.5) |  | 128775 (42.8) | 133966 (44.5) |  |
| Brown/Pardo | 23789 (33.7) | 21471 (30.4) |  | 93042 (30.9) | 82110 (27.3) |  |
| Black/Preta | 3498 ( 5.0) | 2957 ( 4.2) |  | 12804 ( 4.3) | 11176 ( 3.7) |  |
| Yellow/ Amarela | 922 ( 1.3) | 1218 ( 1.7) |  | 4694 ( 1.6) | 5618 ( 1.9) |  |
| Indigenous/Indigena | 197 ( 0.3) | 281 ( 0.4) |  | 399 ( 0.1) | 291 ( 0.1) |  |
| Missing | 13490 (19.1) | 14634 (20.7) |  | 61311 (20.4) | 67864 (22.5) |  |
| **Region of residence** |  |  | <0.001 |  |  | <0.001 |
| North | 3424 ( 4.9) | 3424 ( 4.9) |  | 8355 ( 2.8) | 8355 ( 2.8) |  |
| Northeast | 10216 (14.5) | 10216 (14.5) |  | 50186 (16.7) | 50186 (16.7) |  |
| Central-West | 5758 ( 8.2) | 5758 ( 8.2) |  | 15313 ( 5.1) | 15313 ( 5.1) |  |
| Southeast | 38529 (54.6) | 38529 (54.6) |  | 199212 (66.2) | 199212 (66.2) |  |
| South | 12618 (17.9) | 12618 (17.9) |  | 27959 ( 9.3) | 27959 ( 9.3) |  |
| **Reported number of chronic comorbidities<sup>‡</sup>, n (%)** |  |  | 0.037 |  |  | 0.064 |
| None | 58550 (83.0) | 57585 (81.6) |  | 265890 (88.3) | 271794 (90.3) |  |
| One or two | 11258 (16.0) | 12088 (17.1) |  | 32894 (10.9) | 27565 ( 9.2) |  |
| Three or more | 737 ( 1.0) | 872 ( 1.2) |  | 2241 ( 0.7) | 1666 ( 0.6) |  |
| **Prior SARS-CoV-2 exposure** |  |  |  |  |  |  |
| Previous symptomatic events notified to the surveillance system<sup>¶</sup>, n (%) | 17825 (25.3) | 12137 (17.2) | 0.198 | 88840 (29.5) | 78548 (26.1) | 0.076 |
| Positive SARS-CoV-2 test result<sup>††</sup>, n (%) | 3881 ( 5.5) | 1402 ( 2.0) | 0.186 | 24261 ( 8.1) | 16613 ( 5.5) | 0.101 |
| **Interval between symptoms onset and RT-PCR testing, median (p25-p75), days** | 3 [2, 5] | 3 [2, 5] | 0.115 | 3 [2, 4] | 3 [2, 4] | 0.151 |
| **Hospitalization or Death** | 8097 (11.5) | 12842 (18.2) | 0.191 | 26664 ( 8.9) | 19851 ( 6.6) | 0.085 |
| **Vaccination status** |  |  | 0.294 |  |  | 0.223 |
| Not vaccinated, n (%) | 14100 (20.0) | 19354 (27.4) |  | 31413 (10.4) | 36865 (12.2) |  |
| Single dose, within 0-13 days, n (%) | 676 ( 1.0) | 956 ( 1.4) |  | 301 ( 0.1) | 305 ( 0.1) |  |
| Single dose, ≥14 days, n (%) | 5695 ( 8.1) | 5810 ( 8.2) |  | 14809 ( 4.9) | 14667 ( 4.9) |  |
| Two doses, within 0-13 days, n (%) | 1904 ( 2.7) | 1697 ( 2.4) |  | 796 ( 0.3) | 601 ( 0.2) |  |
| Two doses, 14-89 days, n (%) | 13248 (18.8) | 10877 (15.4) |  | 10758 ( 3.6) | 9808 ( 3.3) |  |
| Two doses, 90-179 days, n (%) | 17605 (25.0) | 17009 (24.1) |  | 84500 (28.1) | 100216 (33.3) |  |
| Two doses, ≥180 days, n (%) | 11431 (16.2) | 12351 (17.5) |  | 30159 (10.0) | 35974 (12.0) |  |
| Third dose of CoronaVac, 0-7 days, n (%) | 68 (0.1) | 54(0.1) |  | 411 ( 0.1) | 368 ( 0.1) |  |
| Third dose of CoronaVac, 8-59 days, n (%) | 328 (0.5) | 271(0.4) |  | 2416 ( 0.8) | 2545 ( 0.8) |  |
| Third dose of CoronaVac, ≥60 days, n (%) | 54(0.1) | 47(0.1) |  | 6412 ( 2.1) | 8309 ( 2.8) |  |
| Third dose of BNT162b2, 0-7 days, n (%) | 929 (1.3) | 837(1.2) |  | 3913 ( 1.3) | 4132 ( 1.4) |  |
| Third dose of BNT162b2, 8-59 days, n (%) | 42421(6.0) | 1187 (17) |  | 29257 ( 9.7) | 17102 ( 5.7) |  |
| Third dose of BNT162b2, ≥60 days, n (%) | 265 (0.4) | 95(0.1) |  | 85880 (28.5) | 70133 (23.3) |  |
| **Interval between first dose and testing, median (p25-p75), days** | 36 [23, 67] | 35 [21, 67] | 0.006 | 138 [93, 170] | 143 [101, 173] | 0.072 |
| **Interval between second dose and testing, median (p25-p75), days** | 144 [60, 181] | 150 [73, 187] | 0.105 | 140 [119, 176] | 142 [122, 178] | 0.039 |
| **Interval between third dose and testing, median (p25-p75), days** | 21 [11, 38] | 10 [6, 30] | 0.307 | 86 [53, 103] | 90 [62, 106] | 0.123 |

RT-PCR=reverse transcription polymerase chain reaction; SMD=standardized mean difference; SD=standard deviation; † Race/skin colour as defined by the Brazilian national census bureau (Instituto Nacional de Geografia e Estatísticas). ‡ Comorbidities included cardiovascular, or renal conditions, diabetes, chronic respiratory disorder, obesity, or immunosuppression. ¶ Reported illness with covid-19 associated symptoms in eSUS and SIVEP-Gripe databases before the start of study on 06 September 2021. \*\* Defined as a positive SARS-CoV-2 RT-PCR or antigen detection test result before the start of study on 06 September 2021.

**eTable 10.** Effectiveness of CoronaVac and homologous or heterologous booster against symptomatic Covid-19 and Covid-19 hospital admission or deaths in adults in Brazil, from the sensitivity analysis including RT-PCR SARS-CoV-2 tests only

|  | Symptomatic COVID-19 |  | Hospitalization or death |  |
| --- | --- | --- | --- | --- |
|  | Delta | Omicron | Delta | Omicron |
| **Not vaccinated** | Reference | Reference | Reference | Reference |
| **Single dose, within 0-13 days** | -11.9% (-20.8--2) | 7.8% (-7.8-21.6) | -16.5% (-49.1-27.1) | - |
| **Single dose, ≥14 days** | 25.5% (22-28.8) | 10.5% (7.9-12.9) | 45.2% (36-53.1) | 47.1% (38.7-54.3) |
| **Two doses, within 0-13 days** | 34.5% (29.3-39.2) | 30.7% (22.7-37.8) | 78.2% (67-85.6) | 67.8% (15.3-87.8) |
| **Two doses, 14-59 days** | 49% (46.8-51.2) | 22.2% (18.2-26) | 85.4% (81.4-88.5) | 23% (-15.7-50) |
| **Two doses, 60-179 days** | 35.5% (33.1-37.8) | -7.1% (-8.9--5.3) | 68.1% (65-71) | 53.4% (47.2-58.9) |
| **Two doses, ≥180 days** | 30.1% (27.3-32.8) | 7% (4.8-9.1) | 57.3% (52.7-61.5) | 55.8% (51.8-59.5) |
| **Third dose of CoronaVac, 0-7 days** | 52.3% (31-67.1) | 18.2% (5.4-29.2) | 76% (57.6-86.4) | 18.4% (-72.3-81.6) |
| **Third dose of CoronaVac, 8-59 days** | 56.9% (47.8-64.4) | 10.5% (5-15.7) | 74.1% (64.5-81.1) | 72.1% (59.7-80.7) |
| **Third dose of CoronaVac, ≥60 days** | 64.1% (41.4-78) | -1.6% (-5.8-2.8) | 84.2% (67.5-92.3) | 63.6% (58.9-67.7) |
| **Third dose of BNT162b2, 0-7 days** | 46.4% (40.4-51.7) | 6.9% (2.3-11.2) | 78.5% (72.5-83.2) | 63.8% (48.8-74.4) |
| **Third dose of BNT162b2, 8-59 days** | 85% (83.8-86.2) | 53.5% (52.3-54.7) | 90.5% (88.7-92) | 84.4% (82.3-86.2) |
| **Third dose of BNT162b2, ≥60 days** | 84.8% (79.9-88.5) | 39.6% (38.4-40.8) | 87.3% (78.1-92.6) | 82.9% (81.4-84.3) |

**eTable 11.** Characteristics of adults in Brazil, who were selected into case-test negative pairs for the sensitivity analysis using RT-PCR SARS-CoV-2 tests only, during the Delta period (September 6, 2021 to December 14, 2021) and Omicron period (December 25, 2021 to April 22, 2022), for the analysis of relative vaccine effectiveness

|  | Matched pairs for Omicron period |  |  |
| --- | --- | --- | --- |
|  | Controls<br>(n=135,053) | Cases<br>(n=135,053) | SMD |
| **Demographics** |  |  |  |
| **Age, mean (SD), years** | 54.1 (19.4) | 54.2 (19.7) | 0.006 |
| **Age categories, n (%)** |  |  | 0.042 |
| 18-39 years | 41810 (31.0) | 41514 (30.7) |  |
| 40-59 years | 34262 (25.4) | 34931 (25.9) |  |
| 60-79 years | 48057 (35.6) | 46309 (34.3) |  |
| ≥80 years | 10924 ( 8.1) | 12299 ( 9.1) |  |
| **Male sex, n (%)** | 46477 (34.4) | 46477 (34.4) | <0.001 |
| **Self-reported race<sup>†</sup>, n (%)** |  |  | 0.126 |
| White/Branca | 61832 (45.8) | 63243 (46.8) |  |
| Brown/Pardo | 38848 (28.8) | 32776 (24.3) |  |
| Black/Preta | 5334 ( 3.9) | 4561 ( 3.4) |  |
| Yellow/ Amarela | 2067 ( 1.5) | 2350 ( 1.7) |  |
| Indigenous/Indigena | 194 ( 0.1) | 176 ( 0.1) |  |
| Missing | 26778 (19.8) | 31947 (23.7) |  |
| **Region of residence** |  |  | <0.001 |
| North | 3859 ( 2.9) | 3859 ( 2.9) |  |
| Northeast | 24618 (18.2) | 24618 (18.2) |  |
| Central-West | 7255 ( 5.4) | 7255 ( 5.4) |  |
| Southeast | 86825 (64.3) | 86825 (64.3) |  |
| South | 12496 ( 9.3) | 12496 ( 9.3) |  |
| **Reported number of chronic comorbidities<sup>‡</sup>, n (%)** |  |  | 0.087 |
| None | 110250 (81.6) | 114600 (84.9) |  |
| One or two | 22967 (17.0) | 19083 (14.1) |  |
| Three or more | 1836 ( 1.4) | 1370 ( 1.0) |  |
| **Prior SARS-CoV-2 exposure \*\*** |  |  |  |
| Previous symptomatic events notified to the surveillance system<sup>¶</sup>, n (%) | 41841 (31.0) | 34182 (25.3) | 0.126 |
| Positive SARS-CoV-2 test result<sup>††</sup>, n (%) | 10419 ( 7.7) | 6303 ( 4.7) | 0.127 |
| **Interval between symptoms onset and RT-PCR testing, median (p25-p75), days** | 3 [2, 4] | 3 [1, 4] | 0.164 |
| **Hospitalization or Death** | 18040 (13.4) | 13484 (10.0) | 0.106 |
| **Vaccination status** |  |  | 0.218 |
| Two doses, ≥180 days, n (%) | 26610 (19.7) | 34817 (25.8) |  |
| Third dose of CoronaVac, 0-7 days, n (%) | 262 ( 0.2) | 357 ( 0.3) |  |
| Third dose of CoronaVac, 8-59 days, n (%) | 1929 ( 1.4) | 2458 ( 1.8) |  |
| Third dose of CoronaVac, 60-89 days, n (%) | 1613 ( 1.2) | 1858 ( 1.4) |  |
| Third dose of CoronaVac, 90-119 days, n (%) | 2694 ( 2.0) | 3811 ( 2.8) |  |
| Third dose of CoronaVac, ≥120 days, n (%) | 1888 ( 1.4) | 2561 ( 1.9) |  |
| Third dose of BNT162b2, 0-7 days, n (%) | 2649 ( 2.0) | 3954 ( 2.9) |  |
| Third dose of BNT162b2, 8-59 days, n (%) | 23637 ( 17.5) | 16503 ( 12.2) |  |
| Third dose of BNT162b2, 60-89 days, n (%) | 26921 ( 19.9) | 23643 ( 17.5) |  |
| Third dose of BNT162b2, 90-119 days, n (%) | 38587 ( 28.6) | 36377 ( 26.9) |  |
| Third dose of BNT162b2, ≥120 days, n (%) | 8263 ( 6.1) | 8714 ( 6.5) |  |
| **Interval between second dose and RT-PCR/Antigen test, mean (SD), days** | 272 [239, 309] | 273 [237, 313] | 0.003 |
| **Interval between third dose and RT-PCR/Antigen test, mean (SD), days** | 88 [57, 104] | 90 [63, 106] | 0.078 |

RT-PCR=reverse transcription polymerase chain reaction; SMD=standardized mean difference; SD=standard deviation; † Race/skin colour as defined by the Brazilian national census bureau (Instituto Nacional de Geografia e Estatísticas). ‡ Comorbidities included cardiovascular, or renal conditions, diabetes, chronic respiratory disorder, obesity, or immunosuppression. \*\* Before the start of study on 06 September 2021. ¶ Reported illness with covid-19 associated symptoms in eSUS and SIVEP-Gripe databases. †† Defined as a positive SARS-CoV-2 RT-PCR or antigen detection test result

**eTable 12.**
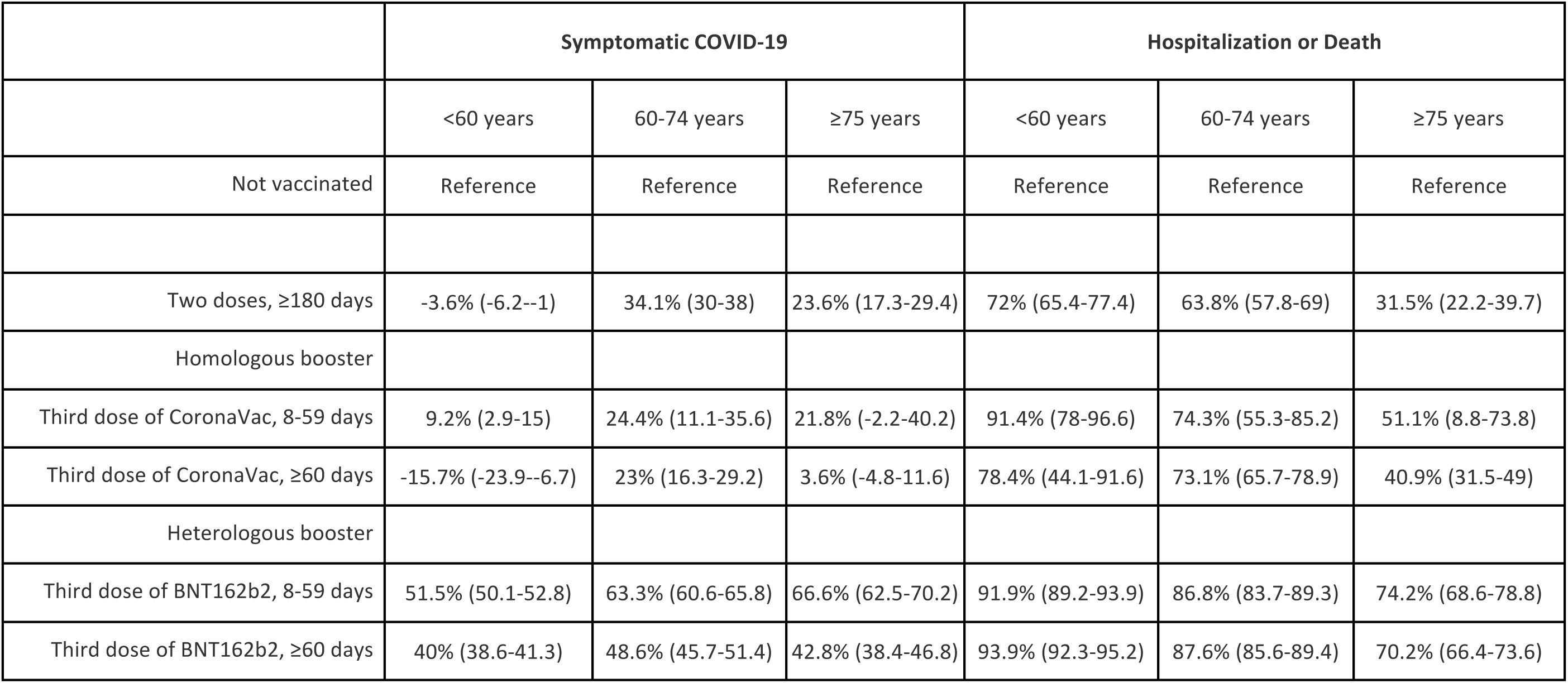
Effectiveness of homologous or heterologous booster against symptomatic Covid-19 and Covid-19 hospital admission or deaths in adults stratified by age during Omicron period in Brazil from the sensitivity analysis including RT-PCR SARS-CoV-2 tests only

**eTable 13.** Vaccine effectiveness of homologous and heterologous booster relative to those at least 180 days after the second dose of a primary series of CoronaVac during the Omicron period, from the sensitivity analysis including RT-PCR SARS-CoV-2 tests only

|  | Symptomatic COVID-19 | Hospitalization or Death |
| --- | --- | --- |
|  | Relative<br>VE (95% CI) | Relative<br>VE (95% CI) |
| Two doses, ≥180 days | Reference | Reference |
| **Homologous booster** |  |  |
| Third dose of CoronaVac, 8-59 days | 1.5% (-5.0-7.7) | 37% (7.5-57.1) |
| Third dose of CoronaVac, 60-79 days | 2.7% (-4.7-9.7) | 34.6% (17.2-48.4) |
| Third dose of CoronaVac, 90-119 days | -19.3% (-24--14.4) | 7.8% (-7.6-21.5) |
| Third dose of CoronaVac, ≥120 days | -20.6% (-26.4--14.3) | 15.9% (3.2-27) |
| **Heterologous booster** |  |  |
| Third dose of BNT162b2, 8-59 days | 49.8% (48.5-51.2) | 62.8% (57.9-67.1) |
| Third dose of BNT162b2, 60-89 days | 35.9% (34.3-37.5) | 68.3% (65.1-71.1) |
| Third dose of BNT162b2, 90-119 days | 30.9% (29.2-32.4) | 62.0% (58.8-64.9) |
| Third dose of BNT162b2, ≥120 days | 16.6% (13.2-19.9) | 50.3% (44.8-55.3) |

